# Dynamic cerebral autoregulation during and 3 months after endovascular treatment in large vessel occlusion stroke

**DOI:** 10.1101/2024.09.05.24313166

**Authors:** Adam Vittrup Heiberg, Troels Gil Lukassen, Thomas Clement Truelsen, Henrik Gutte Borgwardt, Goetz Benndorf, Christine Sølling, Henrik Winther Schytz, Kirsten Møller, Klaus Hansen, Helle Klingenberg Iversen

**Author notes:** Corresponding author, Valdemar Hansens vej 13, N25, DK- 2600 Glostrup, Denmark.

## Abstract

Acute ischemic stroke caused by large vessel occlusion is effectively treated by endovascular treatment (EVT). However, treatment could be further refined by improved understanding of the pathophysiology, including dynamic cerebral autoregulation (dCA). Near-infrared spectroscopy (NIRS) requires virtually no setup time and enables dCA investigation during EVT by measuring dynamic concentration in cortical oxygenated hemoglobin (OxyHb). We aimed to investigate dCA during EVT, before and after recanalization, and at follow-up (FU) after 24 hours and 90 days. We applied interhemispheric transfer function analysis (TFA) of low-frequency (LF) oscillations (0.07-0.2 Hz) in OxyHb that yields the dCA measures, gain and phase shift. LF phase shift increased immediately after recanalization for patients with milder symptom severity and at 24-hour FU for patients with more severe symptoms, but contralateral phase shift needs further investigation. We found higher LF gain in patients with favorable outcome and in patients starting intravenous thrombolysis before EVT. Adjusted for age, infarct size before EVT, and recanalization success, average LF gain predicted independent functional outcome, symptom severity and mortality at 90-day FU. In conclusion, interhemispheric TFA based on NIRS was feasible to assess dCA during EVT and provided insights that could potentially be applied in the development of individualized treatment.

**Clinical Trial Registration:** ClinicalTrial.gov: NCT03738644. https://clinicaltrials.gov/study/NCT03738644.

## Introduction

Stroke remains a worldwide leading cause of disability and death despite the emergence of effective recanalization therapies^1^. Patients with a large vessel occlusions (LVO) face the worst prognosis and constitute up to 38% of patients with acute ischemic stroke (AIS)^2^. Endovascular treatment (EVT) drastically improves functional outcome after anterior circulation LVO when performed within 6 hours^3^, or within 24 hours in cases with mismatch between neurologic deficit and infarct core^4^. Posterior circulation EVT has shown similar advantages compared to best medical treatment^5^. However, procedure-related complications and futile recanalization still restrain the overall efficacy of EVT^6,7^. Impairment of cerebral autoregulation is one of the proposed mechanisms for unexplained futile recanalization^7,8^.

Cerebral autoregulation is the maintenance of suitable blood flow despite changes in cerebral perfusion pressure^9^. Dynamic cerebral autoregulation (dCA) determines how cerebral blood flow is regulated during rapid changes in perfusion pressure^10^. Prevailing methods indicate varying degrees of dCA impairment in patients with AIS including LVO^8,11^ which could endanger vulnerable brain tissue by hypo- and hyperperfusion during and after EVT. Post-thrombectomy studies have shown dCA is an independent predictor of functional outcome^12^, and that it can be used to individualize blood pressure management^13^. However, dCA has never been examined before or during EVT.

Most dCA investigations rely on transcranial Doppler sonography examining blood flow velocity in the middle cerebral artery (V_MCA_) which requires expertise and setup time^14^. Assessment of dCA before EVT would compromise patient safety when every minute counts. Monitoring dCA during EVT also require modalities that do not interfere with the digital subtraction angiography. Near-infrared spectroscopy (NIRS) is an optical method which continuously measures dynamic changes in cortical hemoglobin concentrations without needing any consequential setup time or causing artifacts on the digital subtraction angiography^15,16^.

Low-frequency oscillations (LFO, approximately 0.1 Hz) are prevalent in both systemic (i.e., arterial blood pressure, ABP) and cerebral circulation (e.g., V_MCA_, NIRS)^15,17,18^. Quantifying the changes between them in the frequency domain by transfer function analysis (TFA) is one of the most utilized and standardized methods of assessing dCA^19,20^. TFA quantifies dCA by phase shift and gain. Phase shift is the temporal disruption of LFOs while gain is the amplitude ratio between LFOs. Applying TFA for comparison of the NIRS signal between the unaffected and the ischemic hemisphere, we aimed to investigate dCA before, during and after EVT and associate dCA to index stroke characteristics, treatment and long-term outcome.

## Methods

This prospective observational study (ClinicalTrials.gov: NCT03738644) was conducted between November 2018 and November 2020 after approval from the Scientific Ethics Committees for the Capital Region of Denmark (H-18028704) and in accordance with the World Medical Association Declaration of Helsinki. All participants or their proxy signed written informed consent. Reporting complies with STrengthening the Reporting of OBservational studies in Epidemiology (STROBE) guidelines for observational studies^21^. Corresponding author had full access to all the data in the study and take responsibility for data integrity and analysis. Further methodical details are provided in the supplementary information.

All AIS patients admitted to Rigshospitalet with LVO receiving EVT were screened for eligibility. The exclusion criteria are shown in the enrollment flowchart (Figure 1). Patients with bilateral infarction were excluded as no contralateral hemisphere could be defined (see interhemispheric transfer function analysis section).

**Figure 1.**
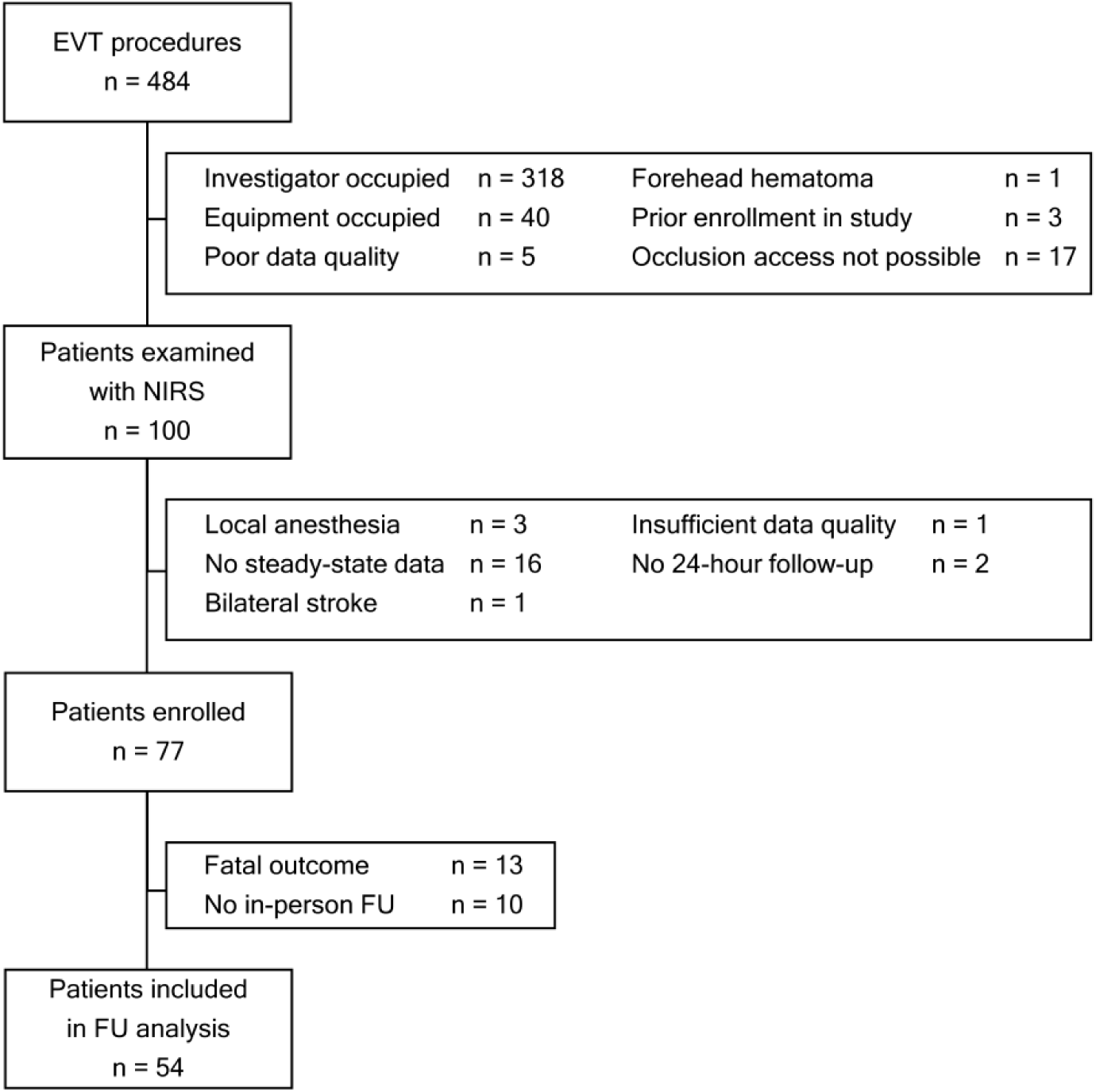
Enrollment and exclusion process. NIRS: Near-infrared spectroscopy. EVT: Endovascular treatment. TFA: Transfer function analysis. FU: Follow-up.

Patients were diagnosed and treated as per standard-of-care. LVO was confirmed and Alberta Stroke Program Early CT Score (ASPECTS)^22^ or Posterior circulation-ASPECTS (PC-ASPECTS)^23^ were determined by staff neuroradiologists assisted by RAPID AI software (iSchemaView, Menlo Park, California, USA) and dichotomized to favorable (ASPECTS/PC-ASPECTS ≥ 6) or non-favorable (ASPECTS/PC-ASPECTS < 6). Patients were transferred immediately to the angiosuite where symptom severity was re-assessed using the National Institutes of Health Stroke Scale (NIHSS) before EVT was performed under general anesthesia without intraarterial vasodilators. In addition to conventional monitoring^24^, NIRS was applied throughout the procedure (described below).

Reperfusion success was assessed by the treating interventional neuroradiologist (modified Treatment In Cerebral Infarction, mTICI; Grade 0-2a unsuccessful achieving no reperfusion to reperfusion in less than half of the occluded artery territory; Grade 2b-3 successful reperfusion in more than half of the occluded artery territory or complete reperfusion)^25^. Post-procedure, the patients were extubated as soon as possible and underwent dual-energy computed tomography (CT) or magnetic resonance imaging (MRI) after 24 hours to detect intracranial hemorrhage or other complications. Patients underwent further examinations as per standard-of-care. Stroke etiology was classified based on the Causative Classification System for Ischemic Stroke^26^.

### Follow-up

Patients had two follow-up (FU) examinations during awareness with NIRS in the supine position for 20 minutes after at least 5 minutes of rest. The first FU was performed 24 hours after EVT (+/- 6 hours from end of endovascular reperfusion efforts) in addition to NIHSS assessment. The second FU was completed after 90 days (+/- 14 days) with functional outcome scored modified Rankin Scale (mRS) and independency (defined as mRS of 0-2) as well as NIHSS. Occurrence of re-hospitalizations, new vascular events (recurrent stroke, myocardial infarction, or surgery for peripheral artery disease) and all-cause mortality were registered. Patients unable to attend 90- day FU, were offered home visits within a 2-hour transportation radius. Patients who did not have in-person FU, were followed up by phone interview and using the electronic health record system.

### NIRS examination

The NIRS monitoring was performed using a continuous wave-NIRS (CW-NIRS) system (Octamon, Artinis Medical Systems, Elst, the Netherlands) with three long-distance channels (35 mm) per hemisphere, examining dynamic hemoglobin concentrations in the prefrontal cortex in the border zones between territories of the middle cerebral (MCA) and the anterior cerebral arteries (ACA) (two channels) and ACA territories exclusively (one channel per side)^27^. A fourth channel with short diode-receiver distance (10 mm) mainly examined the extracerebral tissues on both sides (Figure 2).

**Figure 2.**
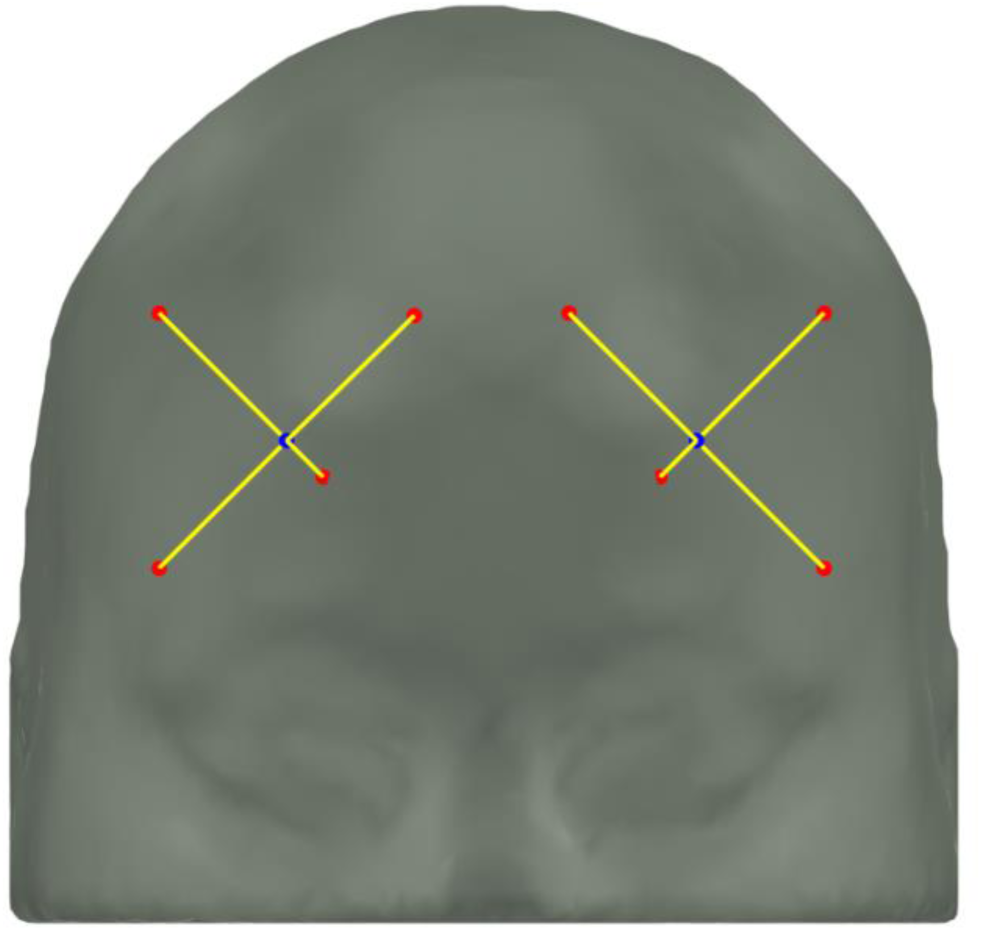
Placement of NIRS channels. Corresponding channels from ischemic and contralateral hemisphere were analyzed by transfer function analysis and averaged subsequently. Short distance-channels were used in separate analysis of extracerebral signals.

### Time segment selection

Per recommendation from the Cerebrovascular Research Network only steady state data segments of five minutes were analyzed^20^. Thus, data segments were excluded within 5 minutes of inducing general anesthesia and within 2 minutes of changes in anesthetics, opioids, or vasopressors^28^. Very noisy data segments were also excluded from the analysis. Individual NIRS signal and all vitals were evaluated for steady state with an allowed variation of 10%. The first data segments free of these exclusionary criteria were chosen.

The first segment (PRE) occurred after sedation but before any attempted revascularization. The second data segment (POST) was just after the final reperfusion status was achieved, including the abandonment of reperfusion efforts. Third (24-hour) and fourth (90-day) data segments were selected based on the extent of motion artifacts. Patients who did not have complete PRE, POST and 24-hour segments were excluded from further analysis.

### Interhemispheric transfer function analysis

The Cerebrovascular Research Network guidelines for TFA^20^ were adapted with minor exceptions described throughout. TFA was only performed on oxygenated hemoglobin (OxyHb) concentrations as they are more reliable^29^ and better reflect the arterial compartment^15^. Input to TFA was OxyHb from the contralateral hemisphere, while the corresponding contralateral channel from the ischemic hemisphere served as output. Fourier transformation was performed, and coherence, gain (amplitude ratio) and phase shift were then calculated in three frequency intervals (Figure 3): High-frequency (HF, 0.2-0.5 Hz), low-frequency (LF, 0.07-0.2 Hz) and very low-frequency (VLF, 0.02-0.07 Hz) and reported as recommended^20^. HF oscillations are transferred passively to cerebral circulation without transformation by dCA. The dCA aspects exhibited in the transformation of VLF oscillations are quite uncertain when limiting time segments to five minutes. Longer time segments were not as attainable in this clinical setting. Therefore, we restricted further analyses to the LF range. Power spectral density (PSD) specifies amplitudes ((µM*mm)^2^/Hz) for each hemisphere.

**Figure 3.**
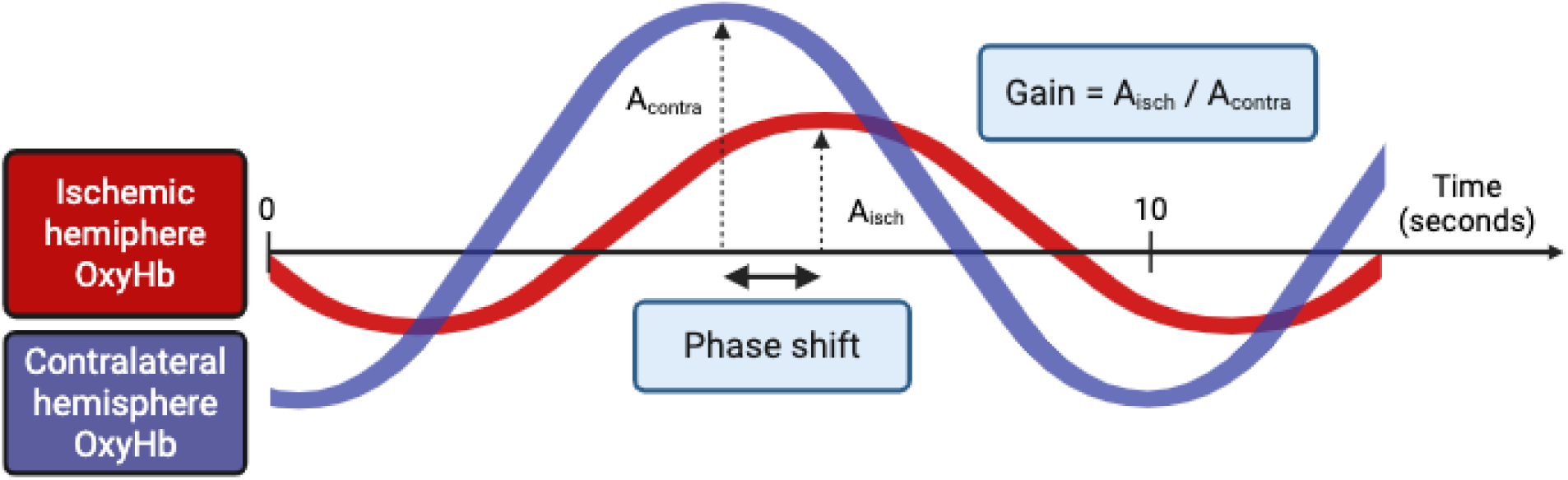
Conceptual illustration of transfer function analysis resulting in phase shift and gain.

Positive phase shifts denoted LFOs in the contralateral hemisphere occurring before LFOs in the ischemic hemisphere and vice versa. Because dyssynchronization of phase shifts can occur in either direction, absolute phase shift was calculated. Similarly, gain of more than 1 resulted from amplitudes in the ischemic hemisphere exceeding amplitudes in the contralateral hemisphere and vice versa. Theoretically, dCA would be deemed intact with no substantial phase shift and gain equal to one. This interpretation assumes dCA in the contralateral hemisphere is intact.

Any differences between input and output signals were assumed to result from intracerebral changes as extracerebral circulation is not under the influence of cerebral autoregulation. We tested this assumption by performing a TFA between short-distance channels from each side with the contralateral side as input and the affected side as output at 24-hour FU, under which the clinical condition was most stable and less prone to artifacts. Segment selection, data preparation and TFA methodology were equal to the processing for long-distance channels described above. Unfortunately, short-distance channels are more prone to oversaturation (i.e., by too much returned infrared light for sensors to record <changes), reducing the number of channels with adequate data quality, especially in acute settings. Thus, incorporation of short-distance channel TFA in statistical models was not possible.

### Statistics

Statistical analysis was performed in two subsets of participants: 1) All participants with sufficient data at PRE, POST and 24-hour FU (FU_24_-group) 2) All patients with sufficient data at PRE, POST, 24-hour and 90-day FU (FU_90_-group) excluding patients with no in-person FU and patients who died. Baseline data from subjects without in-person FU was also analyzed to identify differences in subsets.

Normally distributed data are presented as mean and standard deviation (SD), while non-normal data are presented as median and interquartile range (IQR). Two-sample testing was performed as appropriate. False discovery rate was applied when performing multiple comparisons.

Linear mixed-effect models were fitted with LF gain or LF absolute phase shift as outcome, subjects as random effects, and fixed effects comprised of time segment in addition to and different patient characteristics (e.g., stroke etiology, age, favorable ASPECTS before EVT, anterior or posterior circulation stroke), treatment (e.g., intravenous thrombolysis (IVT), anesthesia, vasopressor, successful recanalization, and occurrence intracranial hemorrhage (ICH) regardless of symptom worsening) or outcome (e.g., NIHSS, mRS, mortality). In case of significant fixed effects, mixed-effect models were fitted with LF PSD from each hemisphere as an outcome to examine the LFO amplitude in each hemisphere. Interaction between time segment and other fixed effects were assessed to determine different temporal developments between groups.

Significant fixed effects from mixed-effect models were used as outcome in logistic regression models with corresponding TFA parameter as univariate predictor. Two multivariate predictions were then performed. The acute prediction model was adjusted for age, recanalization success, and favorable ASPECTS/PC-ASPECTS^30^. NIHSS was not used as co-variate as some patients were intubated before arrival in the angiosuite and did not have a NIHSS before EVT. The 24-hour prediction was modelled with 24-hour NIHSS and age as co-variates. For dichotomous outcomes, receiver operating characteristics curves were generated, and area under the receiver operating characteristic curve (AUROC) was generated to assess the accuracy of the predictor in both univariate and multivariate models without validation subsets.

Models estimates (β) are presented with 95% confidence intervals (CI). Significance level was chosen at 5%.

## Results

### Baseline characteristics

Baseline information and medical history of all enrolled subjects and grouped into subjects with and without in-person 90-day FU are listed in Table 1 and Supplementary Table S1. No differences between the FU and non-FU groups were detected.

**Table 1.** Baseline information and medical history.

|  | All enrolled<br>(n=77) | Follow-up<br>(n=54) | No follow-up<br>(n=23) | P-value |
| --- | --- | --- | --- | --- |
| Age, median (IQR) | 73 (63; 80) | 72 (63.3; 79.8) | 75 (64.5; 80) | 0.446 |
| Female sex, n (%) | 30 (39.0) | 18 (33.3) | 12 (52.2) | 0.135 |
| Caucasian ethnicity, n (%) | 74 (96.1) | 53 (98.1) | 21 (91.3) | 0.211 |
| Right-handed, n (%) | 66 (85.7) | 47 (87.0) | 19 (82.6) | 0.724 |
| BMI, median (IQR) | 26 (23.0; 29.7) | 26.3 (23.3;<br>29.7) | 25.1 (22.0; 28.1) | 0.223 |
| Smoking, n (%) |  |  |  | 0.480 |
| Never | 23 (29.9) | 17 (31.5) | 6 (26.1) |  |
| Former | 29 (37.7) | 18 (33.3) | 11 (47.8) |  |
| Current | 25 (32.5) | 19 (35.2) | 6 (26.1) |  |
| Medical history, n (%) |  |  |  |  |
| Prior AIS | 16 (20.8) | 10 (18.5) | 6 (26.1) | 0.545 |
| Prior TIA | 11 (13.0) | 7 (13.0) | 4 (17.4) | 0.726 |
| Hypertension | 50 (64.9) | 38 (70.4) | 16 (69.6) | 1.000 |
| Diabetes | 10 (13.0) | 6 (11.1) | 4 (17.4) | 0.474 |
| Extracranial artery stenosis ≥ | | | | 0.453 |
| 50% | 33 (42.9) | 25 (46.3) | 8 (34.8) |  |
| Ipsilateral | 30 (39) | 24 (44.4) | 6 (26.1) | 0.201 |
| Contralateral | 27 (35.1) | 16 (29.6) | 2 (8.7) | 0.075 |
| Intracranial artery stenosis | 9 (11.7) | 5 (9.3) | 4 (17.4) | 0.439 |
| Atrial fibrillation | 33 (42.9) | 22 (40.7) | 11 (47.8) | 0.620 |

*BMI: Body-mass index. AIS: Acute ischemic stroke. TIA: Transient ischemic attack.*

Index stroke characteristics, treatment and outcome are listed in Table 2 and Supplementary Table S2. Most of the living patients had in-person FU (82.8%). There were no significant differences concerning etiology, onset, occluded artery, procedural technique, complications, anesthetics, or vasopressors. Subjects who did not complete the 90 day-FU had higher NIHSS before EVT, lower proportion of successful reperfusion and longer time from last-known-well to reperfusion possibly leading to lower 24-hour ASPECTS/PC-ASPECTS as well as higher 24-hour NIHSS, 90-day mRS, and all cause-mortality. Non-FU subjects also experienced a numerically higher occurrence of complications and vascular events. Removing subjects with fatal outcome within 90 days from the non-FU group, did not alter the differences between groups.

**Table 2.** Index stroke, treatment, and outcome.

|  | All enrolled<br>(n=77) | Follow-up<br>(n=54) | No follow-up<br>(n=23) | P-value |
| --- | --- | --- | --- | --- |
| Ischemic hemisphere, right<br>side, n (%) | 44 (57.1) | 32 (59.3) | 12 (52.2) | 0.746 |
| Onset, n (%) |  |  |  | 0.387 |
| Wake-up | 19 (24.7) | 11 (20.4) | 8 (34.8) |  |
| Unwitnessed | 10 (13.0) | 7 (13.0) | 3 (13.0) |  |
| Stroke etiology, n (%) |  |  |  | 0.397 |
| Large-artery atherosclerosis | 28 (36.4) | 21 (38.9) | 7 (30.4) |  |
| Cardio-aortic embolism | 27 (35.1) | 18 (33.3) | 9 (39.1) |  |
| Other causes (dissection) | 5 (6.5) | 2 (3.7) | 3 (13) |  |
| Undetermined causes | 17 (22.1) | 13 (24.1) | 4 (17.4) |  |
| IVT, n (%) | 35 (45.5) | 28 (51.9) | 7 (30.4) | 0.133 |
| Occluded artery, n (%) |  |  |  |  |
| ICA | 6 (7.8) | 4 (7.4) | 2 (8.7) | 1.000 |
| ICA-top | 14 (18.2) | 10 (18.5) | 4 (17.4) | 1.000 |
| ICA-tandem | 12 (15.6) | 11 (20.4) | 1 (4.3) | 0.095 |
| M1 | 50 (64.9) | 36 (66.7) | 14 (60.9) | 0.795 |
| M2 | 23 (29.9) | 17 (31.5) | 6 (26.1) | 0.787 |
| ACA | 8 (10.4) | 5 (9.3) | 3 (13.0) | 0.690 |
| BA | 5 (6.5) | 2 (3.7) | 3 (13.0) | 0.154 |
| PCA | 3 (3.9) | 3 (5.6) | 1 (4.3) | 1.000 |
| ASPECTS / PC-ASPECTS,<br>median (IQR) |  |  |  |  |
| Before EVT | 8 (6; 10) | 9 (7; 10) | 7 (6; 8.5) | 0.081 |
| 24-hour | 7 (5; 8) | 7 (6; 8.75) | 5 (3; 7) | 0.002 |
| Successful reperfusion<br>(mTICI≥2b), n (%) | 69 (89.6) | 51 (94.4) | 18 (78.3) | 0.047 |
| Last-known-well to<br>reperfusion* | 294 (202; 625) | 280 (178; 523) | 344 (283; 887) | 0.033 |
| NIHSS, median (IQR) |  |  |  |  |
| Before EVT | 16.5 (11; 21.25) | 15 (9.25; 20) | 20 (14.25; 23.75) | 0.026 |
| 24-hour | 6 (3; 13.25) | 5 (3; 9) | 16 (8.5; 20.75) | <0.001 |
| 90-day | N/A | 2 (1; 4) | N/A |  |
| 90-day mRS, median (IQR) | 3 (1; 4) | 2 (1; 3) | 6 (4; 6) | <0.001 |
| 90-day independence<sup>†</sup>, n (%) | 36 (46.8) | 32 (59.2) | 4 (16.7) | 0.001 |
| Vascular events<sup>‡</sup>, n (%) | 5 (6.5) | 2 (3.7) | 3 (13) | 0.154 |
| All-cause mortality, n (%) |  |  |  |  |
| 90-day | 13 (16.9) | 0 (0) | 13 (56.5) | <0.001 |
| 1 year | 19 (24.7) | 4 (7.4) | 15 (65.2) | <0.001 |

IVT: Intravenous thrombolysis. ICA: Internal carotid artery. M1: First part of middle cerebral artery. M2: Second part of middle cerebral artery. ACA: Anterior cerebral artery. BA: Basilar artery. PCA: Posterior cerebral artery. ASPECTS: Alberta Stroke Program Early CT Score. PC-ASPECTS: Posterior circulation ASPECTS. mTICI: Modified treatment in cerebral infarction. NIHSS: National Institutes of Health Stroke Scale. EVT: Endovascular treatment. \*: In case of mTICI 0, defined as time of abandoning EVT efforts. †: Modified Rankin Scale 0-2. ‡: Defined as recurrent stroke, myocardial infarction, or surgery for peripheral artery disease at 90-day FU.

### Short distance channels TFA

TFA of short distance channel at 24-hour FU is shown in Supplementary Table S3. Across frequency ranges, gain did not differ from 1 and phase shift did not differ from 0. We found no side-to-side difference for mean OxyHb and PSD.

### Average TFA results

The median time from last-known-well to PRE was 4.0 hours (2.5; 7.9). Mean OxyHb decreased at both POST and 24-hour FU similarly in both hemispheres. PSD increased bilaterally in both VLF and LF range after 24 hours. Overall, vitals were quite steady. TFA showed increased absolute phase difference after 24 hours in the LF range and a temporary reduction after recanalization in the VLF range normalizing after 24 hours. Mean OxyHb, vital parameters, PSD, and the results of the TFA for the FU_24_-group are listed in Supplementary Table S4.

In all ranges, FU subjects showed a PSD reduction between 24 hours and 90 days. Coherence and gain were stable between 24 hours and 90 days. LF and VFL absolute phase differences were reduced between 24 hours and 90 days, while quite stable in HF range. All results for the FU_90_-group are shown in Supplementary Table S5.

## Mixed-effect models

### LF gain

In the FU_24_-group LF gain was significantly lower in subjects with increasing mRS (β=-0.04, CI=[-0.07; -0.02], p=0.002), dependent outcome (β=-0.13, CI=[-0.24; -0.02], p=0.019), increasing NIHSS (β=-1.0%, CI=[-1.8%; -0.1%], p=0.029), and in subjects with fatal 90-day outcome (β=-0.21, CI=[-0.35;-0.06], p=0.005) across PRE, POST and 24-hour data segments. The observed reduction in LF gain was the result of greater PSD reduction in the ischemic hemisphere. Patients treated with IVT had higher LF gain (β=0.22 CI=[0.12; 0.32], p<0.001). Treatment with IVT is contraindicated with known onset > 4.5 hours and extensive infarction changes on CT or MRI and could confound the effect of IVT. Therefore, we accounted for time from last-known-well to arrival or ASPECTS/PC-ASPECTS but the IVT influence on LF gain remained significant (Table S6). The effect of mRS and IVT was independent of each other in multivariate model including both variables (IVT: β=0.19, CI=[0.09; 0.29], p=0.001; 90-day mRS: β=-0.03, CI=[-0.06; -0.00], p=0.041).

Grouping patients by anesthetics, vasopressor, stroke etiology (large-artery atherosclerosis or cardioembolic), age, occlusion complication level (simple isolated thrombus or complicated ICA-top or tandem thrombus), favorable ASPECTS/PC-ASPECTS before EVT, anterior or posterior circulation stroke, occurrence of ICH, and hypotension during recording (average mean arterial pressure (MAP) < 70 mmHg) had no effect on LF gain. Grouping patients by recanalization success (unsuccessful: β=-0.15, CI=[-0.33; 0.03] , p=0.105) and hypertension (MAP< 130 mmHg before recanalization or 90 mmHg after recanalization) during recording (β=0.09, CI=[-0.01; 0.19], p=0.082) showed no statistical change in LF gain. There was no interaction effect between the time segment and any other fixed effects. Time segment had no effect on LF gain with PRE as index.

Associations between LF gain and outcome were not prevalent in the FU_90_ group. While LF gain was still numerically lower in subjects with worse outcomes, the difference did not reach a significance level. The association between LF gain and IVT remained in the FU_90_ group (β=0.16, CI=[0.07;0.26], p=0.011). In contrast to FU_24_-group a significant interaction effect between favorable ASPECTS/PC-ASPECTS and the time segment was observed (p=0.035) as LF gain increased more after recanalization and declined more at 24-hour and 90-day FU in patients with non-favorable ASPECTS/PC-ASPECTS. The interaction effect is plotted in Supplementary Figure S1. LF gain was numerically lower at 24-hour and 90-day FU compared to PRE and significantly lower compared to POST.

### LF phase shift

Mixed-effect models showed no effect on absolute phase shift of grouping patients by mRS, mortality, vasopressor, stroke etiology, age, occlusion complication level, favorable ASPECTS/PC-ASPECTS before EVT, anterior or posterior circulation stroke, occurrence of ICH, hypotension nor hypertension during recording. Compared to patients anesthetized with propofol, those receiving sevoflurane showed a trend towards numerically lower absolute phase shift (β=-46.0%, CI=[-71.9%; +3.9%], p=0.064). We found an interaction between NIHSS and time segment as subjects with milder stroke severity had an immediate increase in absolute phase shift after recanalization, as opposed to patients with greater stroke severity increasing in absolute phase shift between POST and 24-hour FU (Figure 4).

**Figure 4.**
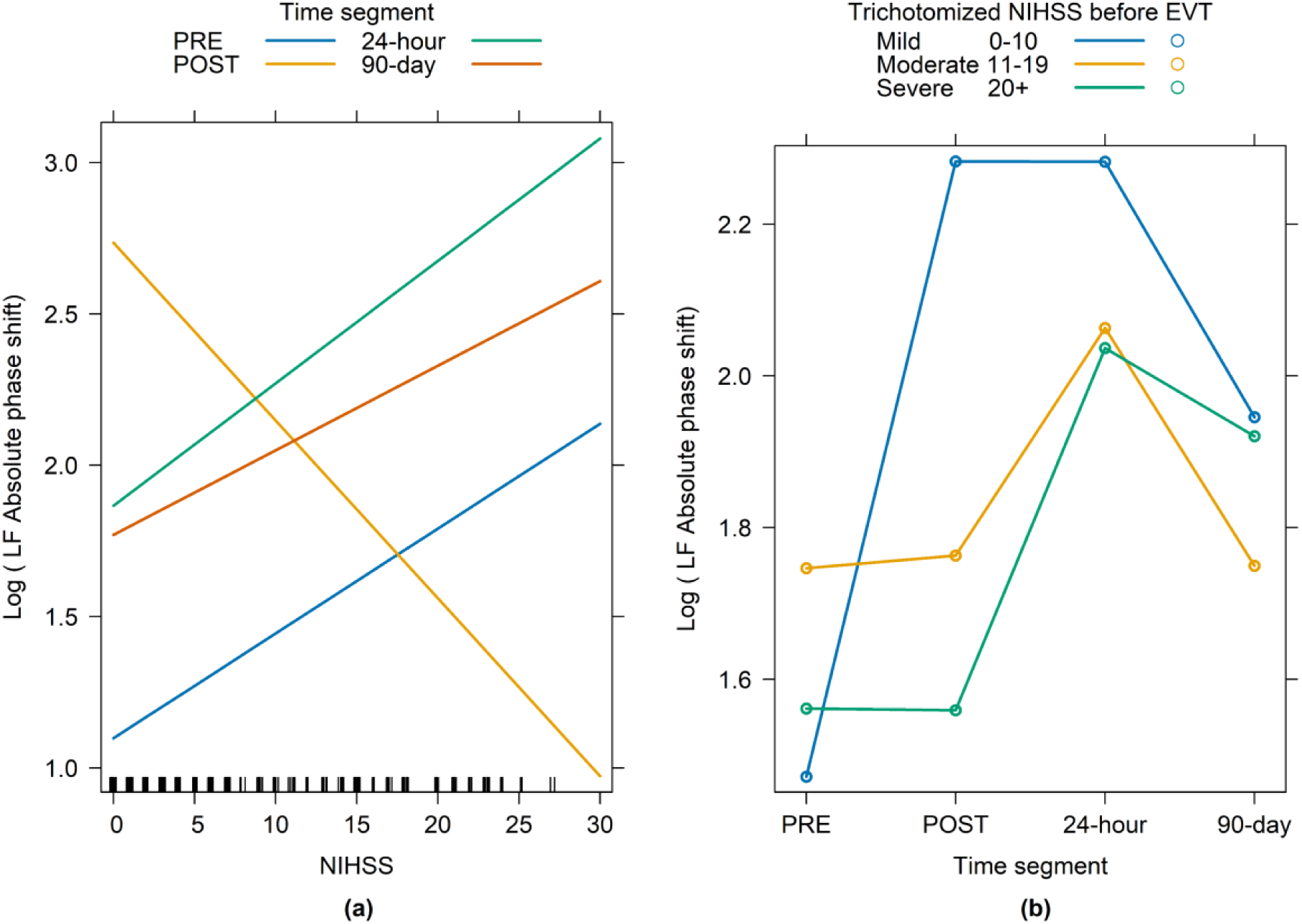
Interaction between time segment and NIHSS in 90-day subsets. (a) Shown as continuous NIHSS (actual model). Non-parallel regression lines indicate interaction clearly seen for POST. (b) Trichotomized NIHSS before EVT (approximation model to illustrate interaction over time segments).

Results from 24-hour and 90-day subsets was equivalent. Absolute phase shift decreased between 24-hour and 90-day FU and was not different from index (PRE) at 90-day FU.

All results from linear mixed-effect models can be found in Supplementary Tables S6-S9.

### Prediction models

LF gain showed associations to outcome and therefore, was used as co-variate in the following prediction models. LF gain was averaged across PRE, POST, and 24-hour FU because mixed-effects models showed no effect of time segment on LF gain. Thus, average LF gain was representable for LF gain in the first 24 hours. It could theoretically still be used to predict outcome in the large proportion of enrolled subjects excluded from analysis due to missing state period (16%). The full output from multivariate models is reported in the supplementary information.

### Modified Rankin scale

Logistic regression models were used to predict 90-day independent (mRS 0-2) or dependent (mRS > 2) functional outcome with LF gain as predicting variable. Average LF gain was significant predictor of independent outcome (β=2.36, CI=[0.41; 4.53], p=0.023, AUROC=0.641) with an odds ratio (OR) of 1.60 (CI=[1.07;2.41]) for a 0.2 increase in LF gain.

In the acute predictions model (adjusted for age, recanalization, and favorable ASPECTS/PC-ASPECTS before EVT), average LF gain remained a significant predictor of independent outcome (β=2.60, CI=[0.32; 5.22], p=0.035, AUROC=0.802, Supplementary Table S10) with an OR of 1.68 (CI=[1.04; 2.73]) for a 0.2 increase in LF gain. Age was the only other significant predictor (p=0.001).

The 24-hour model adjusted for age and trichotomized 24-hour NIHSS (0-5; 6-13; >14) and showed that LF gain (β=2.29, CI=[-0.14; 5.11], p=0.081, AUROC=0.876, Supplementary Table S11) had an OR of 1.58 (CI=[0.94; 2.65] for 90-day independency with a 0.2 increment. Age (p=0.002) and trichotomized 24-hour NIHSS (p=0.001) were significant predictors.

### NIHSS

Predicting 1-step changes in 90-day NIHSS is of minor clinical importance prompting NIHSS categorization (Near-remission 0-1; Mild 2-3; Moderate 4-6; Severe 7-18; Fatal outcome). Ordinal logistic regression model was used to predict 90-day categorized NIHSS with average LF gain as a significant predictor (β=2.14, CI=[0.42; 3.94], p=0.016). With an increase of 0.2 in LF gain, the common OR was 1.53 (CI=[1.08;2.17]) for better categorical 90-day NIHSS. Predicted probabilities for the univariate model is depicted in Figure 5.

**Figure 5.**
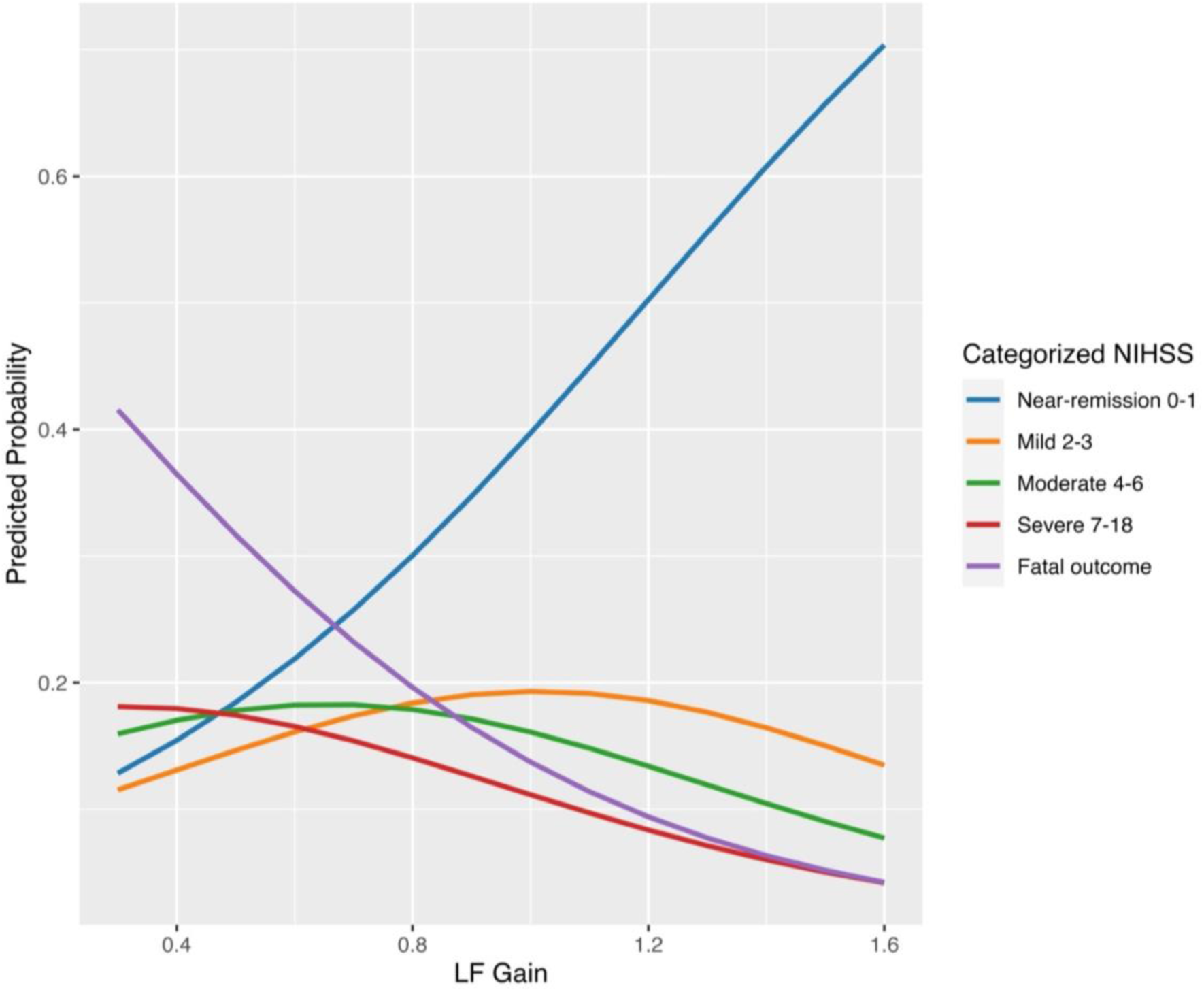
Predicted probabilities of initial LF gain on categorized NIHSS.

In multivariate acute prediction, LF gain remained a significant predictor (β=2.26, CI=[0.45; 4.19], p=0.017, Supplementary Table S12) with a common OR of 1.57 (CI=[1.08; 2.28]) for better categorical 90-day NIHSS. Favorable ASPECTS/PC-ASPECTS before EVT was the only other significant predictor (p=0.015).

The 24-hour model showed average LF gain (β=3.11, CI=[-0.46; 3.66], p=0.072, AUROC=0.876, Supplementary Table S13) had an OR of 1.37 (CI=[0.91; 2.07] for better categorical 90-day NIHSS with a 0.2 increment. Trichotomized 24-hour NIHSS was highly predictive for 90-day NIHSS (p<0.001).

### Mortality

Logistic regression models were used to predict 90-day survival. In the univariate model, average LF gain was a significant predictor of 90-day survival (β=3.73, CI=[1.04; 6.85], p=0.011, AUROC=0.716) with an OR of 2.11 (CI=[1.19; 3.73]) for a0.2 increase in LF gain. In the acute model, average LF gain remained a significant predictor of 90-day survival (β=3.78, CI=[0.77; 7.35], p=0.022, AUROC=0.863, Supplementary Table S14) with an OR of 2.13 (CI=[1.12; 4.06]) for a 0.2 increase in LF gain. Favorable ASPECTS/PC-ASPECTS before EVT was the only other significant predictor (p=0.013).

The 24-hour model (Supplementary Table S15) was strongly affected by collinearity between trichotomized NIHSS and LF gain.

## Discussion

Dynamic CA (dCA) in AIS is of immense interest due to its pathophysiological role and consistent association with long-term outcome^8,31^. Further, dCA has shown promising potential for individualized blood pressure management after thrombectomy^13^. In order to investigate the potential for individualizing intraprocedural treatment it is important to study dCA in the hyperacute phase of stroke and LVO before, during and after revascularization. By our account, this study is the first to examine dCA not only during EVT but also before and after recanalization and possibly the earliest dCA assessment after ischemic stroke. Here, we have shown the time course of interhemispheric TFA based on cortical OxyHb across hyperacute, acute, and chronic phases of LVO and that higher LF gain was associated with IVT prior to EVT and good 90-day functional outcome independent of each other. LF gain also exhibited predictive capabilities of 90- day symptom severity, functional outcome, and mortality.

### Phase shift

Studies of dCA in AIS patients have predominantly been based on transcranial Doppler sonography and TFA between ABP and V_MCA_. Associations between impaired dCA in the ischemic hemisphere and poor outcome have been shown in numerous studies regarding infarct size, edema, hemorrhagic transformation, symptom severity at discharge and long-term functional outcome (mRS) but mostly with LF phase shift as covariate^8,32^.

While we did not find associations between interhemispheric phase shift and functional outcome, our data did show an interaction between NIHSS and time segments. Absolute phase shift increased immediately after recanalization in patients with milder symptom severity whereas patients with higher NIHSS exhibited an increase at 24-hour FU. The differences in temporal development of phase shift dyssynchronization could be related to the penumbra as there were no effect of ASPECTS/PC-ASPECTS nor the recanalization success. Dichotomizing recanalization to complete (mTICI 3) or incomplete (mTICI 2a or 2b), Sheriff et al. also showed an interaction effect when examining ABP-V_MCA_ phase shift over time^33^. Such interactions emphasize the importance of examination time and the need to account for clinical characteristics when studying dCA in AIS patients. Phase shift between ABP and V_MCA_ have shown no interhemispheric difference within 6 hours of onset with moderate to severe stroke symptoms in the MCA territory^34^. Later, reductions compared to healthy controls and contralateral hemisphere are consistently seen from 20 hours and throughout 7 days from onset in patients with MCA occlusion^12,35,36^, while perhaps already normalizing after 10 days^35^.

Using interhemispheric TFA, we found an absolute phase shift relatively close to 0, indicating equal dCA across hemispheres. Studies of the contralateral hemisphere in LVO have shown inconsistent results concerning dCA impairment. Contralateral phase shift dyssynchronization could be the results of vasoactive substances accumulating^7,8^ and perhaps explain why we found no association between interhemispheric phase shift and functional outcome despite consistent association reported to ABP-V_MCA_ phase shift in the ischemic hemisphere^8^. Examining bilateral ABP-V_MCA_ phase shift and interhemispheric phase shift concurrently could reveal this in the future. In this study, simultaneous interhemispheric TFA and bilateral ABP-OxyHb TFA would have diminished the sample size greatly as many patients did not have sufficient ABP segments before recanalization.

Only one study has previously used OxyHb as both TFA input and output in AIS patients^37^. This study was performed 1-2 days after onset and showed increased LF absolute phase shift in non-thrombolyzed patients compared to thrombolyzed patients. The thrombolyzed patients had a higher NIHSS at admission, but a higher rate of total remission and conceivably less infarction volume, perhaps resulting in equal dCA across both hemispheres. The current study found no such difference as effect of recanalization success or IVT probably because of substantial differences concerning study design, cohort and TFA methodology.

### Gain

Similar to the current study, Castro et al. showed that LF gain between ABP and V_MCA_ in the affected hemisphere of moderate to severe MCA strokes within 6 hours of onset was a predictor of 90-day dependency^38^. Gain remained stable between hyperacute (<6 hours), acute (24 hours) and chronic (90-day) stroke stages^38^, while inconsistent findings are reported in the subacute stage (3 to 7 days after onset)^33,39^. However, gain in the ischemic hemisphere increased with outcome severity, interpreted as an increasingly impaired dCA with a diminished ability to dampen the amplitude of LFOs from systemic perfusion^40^. Conversely, a recent meta-analysis found lower gain and thus more intact dCA across all AIS patients compared to healthy controls^11^. The inconsistencies concerning ABP-V_MCA_ gain could be explained by heterogeneities between stroke populations and methodical variability including scaling methods that is required when using different modalities in TFA. LF gain in patients with favorable outcome from the current cohort was close to one, indicating equal dCA between hemispheres and intact dCA in the ischemic hemisphere if intact contralateral dCA is assumed. With increasing outcome severity, LFO amplitude was relatively stable in the contralateral hemisphere while LFO amplitude in the ischemic hemisphere declined resulting in lower gain. The results indicate favorability of increased LFO amplitude and higher gain towards long-term outcome. Favorable outcome is usually associated with intact dCA which in the conventional interpretation of ABP-V_MCA_ gain would be the result of reduced LFO amplitude and lower gain. Thus, direct comparisons of LF gain between conventional TFA setup (ABP-V_MCA_) and interhemispheric TFA seems implausible perhaps due to differences in the examined vasculature. NIRS should be sensitive to all microvascular regulations, while that may not be true for V_MCA_, which is based on upstream arteriolar resistance^41^ and susceptible to changes in vessel diameter^42–44^. Multiple TFAs based on simultaneous measurements of ABP, V_MCA_ and OxyHb would be needed in the future to determine how different TFA models interact.

Therefore, we can only compare amplitude and gain directly to subacute patients that Phillip et al. examined^45^. Numerically amplitude in the ischemic hemisphere was higher resulting in higher gain in IVT patients while neither reached statistical significance^37^. This seems consistent with the current study that showed a substantial effect of IVT but also with a large study based on TCD that found better dCA in IVT patients^46^. The effect of IVT was independent from 90-day mRS and was not confounded by time from last-known-well nor structural damage determined by ASPECTS/PC-ASPECTS. Other possible confounders including current anticoagulant treatment, uncontrollable hypertension, co-morbidities, recent stroke or bleedings was not accounted for, but the results indicate a direct and positive effect of IVT on dCA. The influence does not seem to be a pharmacological effect of IVT but rather the effect of recanalization^46,47^. In LVO patients IVT can impact recanalization both before and after EVT with superior outcome compared to EVT alone^48^. We found no statistical effect of recanalization, but we did not discriminate between levels of successful recanalization (mTICI 2b and 3) nor account for collateral circulation and possible reocclusions after EVT in the recanalization evaluation.

LF gain was a significant predictor of 90-day categorized NIHSS, functional independency and mortality after adjusting for age, recanalization success, and favorable ASPECTS/PC-ASPECTS in the acute prediction model. The 90-day NIHSS categorization was distinguished by relatively small differences in the milder categories but could represent larger differences in lesion size which could explain the large predictive differences between categories. With the added knowledge in the 24-hour prediction model, trichotomized NIHSS seemed to confound the effect of average LF gain on predictions of independency and categorized NIHSS at 90-day FU. Average LF gain could still be impactful in the 24-hour prediction model especially when 24-hour NIHSS is unknown (e.g., due to prolonged intubation).

### Strengths and limitations

In this study, we applied one of the most frequently used and standardized methods to evaluate dCA, though with the novelty adjustment of using NIRS signals as both input and output. There are certain advantages to this approach. This includes easy and fast setup in the hyperacute setting, and a very low proportion of excluded patients due to inadequate signal quality. Further, the applied model accounts for extracerebral tissue contamination because LFO in the extracerebral tissue can reasonably be assumed to be static and without regional differences which was upheld in the short-distance channel TFA. Using NIRS we found absolute certainty of intracerebral signal component due to contrast artifacts in all long-distance channels. Thus, we are confident that any changes in LF gain or phase shift are due to intracerebral changes, i.e. dynamic autoregulation.

Previous studies have attempted other TFA setups to examine dCA based on NIRS alone^49–51^. Such approaches benefit from the ability to perform unilateral dCA assessments but were reliant on either sufficient data quality of short-distance channels or deoxygenated hemoglobin which are more prone to poor data quality. All dCA assessments based on NIRS including the current study will be restricted by the low spatial resolution limiting assessment to the superficial parts of the cerebral cortex. Deeper parts of the brain are equally affected by LVO but whether the observed dCA changes extent into such regions cannot be determined with NIRS.

Various limitations restrict the conclusions from this study. Interhemispheric TFA examines dCA relative to the contralateral hemisphere. While amplitude can be assessed per hemisphere (PSD) phase shift cannot. This leaves uncertainty as to whether phase shift is equally impaired or equally intact in patients with LVO when not doing simultaneous TFA between ABP and OxyHb on each hemisphere. Also, we had no control group of healthy subjects to compare with.

AIS patients including those receiving EVT are inherently heterogenous concerning stroke etiology, co-morbidities, imaging parameters, and treatment. In this study we chose to prioritize everyday generalizability and not exclude patients based on heterogeneities, but account for as many of them as possible in the statistical models instead. Examples of this approach is our analyses of posterior circulation LVO, sevoflurane anesthetics and ICH occurrence which did not influence dCA and therefore did not lead to exclusion of such patients. However, accounting for all heterogeneities is nearly impossible and requires a very large sample size especially for less common occurrences. Unidentified confounders could be affecting the clinical course and the progression of dCA between EVT and follow-up examinations. Unexamined co-morbidities, complications related to EVT or index stroke, rehabilitation efforts, and new vascular events including reocclusions could be possible confounders which would need to be investigated in future studies.

As we considered the study to be exploratory and hypothesis generating, we did not prespecify a statistical analysis plan. Thus, the presented findings deserve confirmation in larger cohorts with prespecified analysis.

The main exclusion criteria from patients examined with NIRS was the absence of a steady state period mainly before recanalization. Reasons for this were a short procedural time to access the occlusion and attempt recanalization within exclusion period after intubation, difficulties to establish steady state, e.g. due to cardiovascular comorbidity as well as high or low sensitivity to anesthetics or vasopressors. Therefore, we believe no selection bias has been introduced by data segment selection.

We did not evaluate collateral circulation due to differences in standard-of-care imaging (CT-angiography, MRI time-of-flight angiography) and missing contralateral series from the digital subtraction angiography. Collateral circulation could certainly have an impact on both outcome and dCA^52,53^. However, collaterals are better developed in patients with large-artery stenosis compared to patients with cardioembolic stroke etiology and we did not find any effect of etiology.

Results from the FU_90_-group were affected by attrition bias as non-FU patients did not attend mainly due to very poor functional or fatal outcome and the results should therefore be interpreted with caution. Efforts were made to minimize this bias by offering FU visits at the rehabilitation center or at home visits despite COVID-19 restrictions. The exclusion of a considerable portion of patients with the worst outcome may explain why associations between LF gain and outcome were not significant in the FU_90_-group.

In conclusion, interhemispheric TFA based on NIRS is a feasible method for investigating dCA in AIS patients before, during and after EVT. Development of LF phase shifts depended on symptom severity and increased immediately after recanalization in AIS patient with lower NIHSS but concurrent dCA investigations using different modalities are warranted to assess dCA in healthy subjects and in the contralateral hemisphere to understand the interplay between different dCA models. LF gain remained stable during EVT up to 90 days post-treatment and had associations to IVT treatment and long-term outcomes of symptom severity, functional outcome, and mortality. Further, LF gain significantly predicted such long-term outcomes. Future studies should confirm these exploratory findings but the study indicates a potential for individualized treatment.

## Data Availability

All processing code is open source available. NIRS data segments can be provided upon reasonable request. Individual patient data are identifiable, and availability requires approval of both centers as well as local ethics committees.

## Non-standard Abbreviations and Acronyms

ACA: Anterior Cerebral Artery
AIS: Acute Ischemic Stroke
ASPECTS: Alberta Stroke Program Early CT Score
BA: Basilar artery.
CW-NIRS: Continuous Wave Near-Infrared Spectroscopy
dCA: Dynamic Cerebral Autoregulation
EVT: Endovascular Treatment
FU: Follow-up
FU_24_-group: All participants with sufficient data at PRE, POST and 24-hour FU.
FU_90_-group: All patients with sufficient data at PRE, POST, 24-hour and 90-day FU.
HF: High-frequency (0.2-0.5 Hz)
ICA: Internal carotid artery.
IVT: Intravenous thrombolysis.
LF: Low-frequency (0.07-0.2 Hz)
LFO: Low-frequency oscillations
LVO: Large vessel Occlusion
M1: First part of middle cerebral artery.
M2: Second part of middle cerebral artery.
MCA: Middle Cerebral Artery
mRS: modified Rankin Scale
mTICI: Modified Treatment In Cerebral Infarction
NIHSS: National Institutes of Health Stroke Scale
NIRS: Near-Infrared Spectroscopy
OxyHb: Oxygenated Hemoglobin Concentration
PC-ASPECTS: Posterior circulation Alberta Stroke Program Early CT Score
PCA: Posterior cerebral artery.
POST: Second time segment (after achieving final reperfusion status)
PRE: First time segment (after sedation but before any attempted revascularization)
PSD: Power spectral
STROBE: STrengthening the Reporting of OBservational studies in Epidemiology
TFA: Transfer Function Analysis
TIA: Transient ischemic attack
VLF: Very low-frequency (0.02-0.07 Hz).
VMCA: Flow velocity in the Middle Cerebral Artery

## Author contributions

A.V.H. and H.K.I. conceived and designed the study with contributions from T.C.T., H.G.B., G.B., C.S., and H.W.S. A.V.H. acquired the data with major contributions from T.G.L. as well as T.C.T., H.G.B., G.B., C.S. and K.H. A.V.H. performed data analysis with contributions from T.G.L. and H.K.I. All authors contributed to interpretation of results. A.V.H. drafted the article. All authors revised the article critically and approved the final version. All persons designated as authors qualify for authorship and agree to be accountable for all aspects of the work and resolve of questions related to the accuracy or integrity of the study.

## Data availability statement

All processing code is open source available at https://openfnirs.org/software/homer/ for preprocessing and at https://www.car-net.org/tools for TFA. NIRS data segments can be provided upon reasonable request. Individual patient data are identifiable, and availability requires approval of both centers as well as local ethics committees. All data requests can be made by contacting the corresponding author.

## Additional Information

The authors declare that there are no competing interests.

