## Supplementary information for "Dynamic cerebral autoregulation during and 3 months after endovascular treatment in large vessel occlusion stroke"

### Manuscript title

### Supplementary Methods

*Medical history*

Baseline diagnosis of hypertension was based only on regular use of antihypertensive medication. Dyslipidemia diagnosis was based on regular use of cholesterol-lowering medication or Low-Density Lipoprotein > 3.0 mmol/L. Diabetes diagnosis was based on regular use of antidiabetic medication or consecutive Hemoglobin A1c measurements above 48 mmol/mol.

*Imaging*

Alberta Stroke Program Early CT Score (ASPECTS)[1] or Posterior Circulation-ASPECTS (PC-ASPECTS)[2] were evaluated based on standard-of-care imaging using highest precision modality by the order magnetic resonance imaging (MRI), computed tomography-perfusion (CTP), and non-contrast computed tomography (NCCT).

*Anesthesia*

Patients were put under general anesthesia maintained by propofol or sevoflurane as well as remifentanil with phenylephrine or norepinephrine as vasopressor infusion. Patients were monitored by NIRS, invasive blood pressure, peripheral pulse oximetry and 3-lead electrocardiogram (Philips IntelliVue, Philips Medical Systems, Eindhoven, The Netherlands) from arrival with vitals being exported by VSCapture.[3] Mechanical ventilation (Dräger, Lübeck, Germany) was adjusted by inspiratory and expiratory levels of $O_2$ and $CO_2$ as well as vitals. Ventilation parameters were exported directly to the electronic health record system at average values per minute. For dichotomizing hypotension during recording was defined as mean arterial pressure (MAP) < 70 mmHg on average,[4] while hypertension during recording was defined as MAP > 130 mmHg before recanalization and MAP > 90 mmHg after recanalization.[4] Intravenous labetalol was administered per standard of care to avoid hyperperfusion (ABP > 185/105 mmHg).[5]

*NIRS examination*

The Octamon system (Artinis Medical Systems, Elst, the Netherlands) consists of dual-wavelength diodes (750 and 839 nm) paired with ambient light-protected receivers relayed to integrative computer by Bluetooth connection. The optodes was arranged in a headband and placed approximately 1 cm over the glabella with lateral projections just superior to the auricular helices. Digital spatial registration was not possible in the hyperacute setting as it would have delayed EVT. To minimize motion artefacts (MA) and avoid optode repositioning the headband was wrapped firmly with adherent gauze wrap. Bluetooth receiver was re-positioned to placement above the vertex to avoid artefacts in the preferred projections of the digital subtraction angiography (DSA). Reflected light intensity was sampled at 25 Hz using OxySoft version 3.0.103.3 (Artinis Medical Systems, Elst, the Netherlands) and exported for further processing and analysis. While not in agreement with CARNet recommendations, the sampling frequency still complies with the Nyquist sampling theorem. Furthermore, the CARNet study showed no difference in ABP-$V_{MCA}$ gain and phase between sampling frequencies 50, 20 and 10 Hz in both LF range and for gain in the VLF range.[6] However, no similar studies have not been done using NIRS.

*Data preparation*

Raw light intensity was quality inspected channel-by-channel to prune any channels without clear cardiac pulse waves. Next artefacts from intracerebral contrast injection were identified to confirm an intracerebral component in the NIRS signal. Intensity was then converted to optical density (OD) before performing correction of smaller (maximum 3 heart beats in coherence with CARNet guidelines[7]) motion artefacts (MA) by the movement artefact reduction algorithm (MARA)[8] after individual identification applying the threshold of two times the average beat-to-beat amplitude in dubious cases. Automatic MA identification with identical parameters across subjects is preferred in fNIRS studies to avoid investigator bias, but the drawback is unidentified artefacts as parameters seldomly suit all recordings. Thus, manual identification was preferred in this study as even a few artefacts could alter the results of the transfer function analysis (TFA)[9] and all recordings were preprocessed before proceeding to TFA to avoid bias. MARA corrections are well-suited for removing baseline shifts but can sometimes be inadequate in removing spikes. Thus, after converting OD to hemoglobin concentrations by the modified Beer-Lambert law (MBLL), concentration data was run through a 3$^{rd}$ order median filter. Channels that could not be corrected adequately were excluded from further analysis. If two out of three corresponding channels did not pass data quality inspection, the subject was excluded. To avoid spectral leakage, the mean of each channel was subtracted from the concentration signals. OxyHb data was low-pass filtered with a third-order Butterworth filter cut off at 0.2 Hz to eliminate high-frequency components. While the majority of TFA studies of CA employ beat-to-beat-averaging and subsequent resampling, low-pass filtering is an equally well-suited method when data is free of artefacts, which is true after careful data selection, pruning of noisy channels and recordings, and MA correction.[10] Using the Welch' method the fast Fourier transform was applied in 100-second Hanning-windows with 50% overlap resulting in 5 windows across the data segments of 5 minutes. Triangular moving average window was applied for spectral smoothing. Coherence, gain (amplitude ratio), and phase shift were then calculated in the HF, LF and VLF frequency intervals as is recommended when analyzing spontaneous oscillations that have some individual variation[7,11,12]. Coherence is used as a quality control as TFA results is rejected when coherence is lower than the threshold corresponding to the number of windows in the recording.[7] As input and output signals should be relatively tightly synchronized, negative phase shifts were not removed from analysis as is recommended with an expected positive phase shift (e.g. TFA between ABP and $V_{MCA}$)[7]. Preliminary analysis of differences between channels showed no effect of channel placement and included channels were therefor averaged. All data processing was performed in MATLAB version R2018b (The MathWorks Inc., Natick, Massachusetts). Preprocessing from light intensity to OxyHb was performed with HomeR package version 2.8[13] while TFA was performed with CARNet package.[7]

*Statistics*

All statistical analysis was performed in R 4.3.3 (R Core Team 2023, Vienna, Austria).

Power calculations were aimed to detect differences in interhemispheric gain and phase shift between patient with independent and dependent 90-day outcome. Significance level was set at 0.05 and power at 0.80. The only study of interhemispheric gain was performed on subacute stroke patients having received thrombolysis or not[14]. Based on an equal effect size and a pooled and weighted standard deviation the required sample size was 78. The effect size was deemed conservative as the clinical difference would surely be greater in a cohort of EVT patients. Interhemispheric phase shift in large vessel occlusions was estimated from two studies. Based on the smallest effect size and the largest variance the required sample size was 50. Accounting for exclusion based on data quality and available steady state segments we aimed for a sample size of 100 patients examined with NIRS.

Testing of independent samples were performed with Welch' t-test or Wilcoxon-Mann Whitney test in case of non-normality, while dependent populations samples were tested with paired t-test or Wilcoxon signed ranks test for non-normal data. Normality was assessed visually by histograms and quantile-quantile plots. Categorical proportions were tested with chi-squared test or Fisher's exact test depending on the number of categories.

In mixed-effect models, compound symmetry was chosen as correlation matrix to avoid overestimation of correlations between recordings just before and after reperfusion, while still maintaining the order of recordings.

Significant interaction between fixed effects rendered reporting of isolated fixed effects meaningless. Insignificant interactions were omitted from models before estimation of fixed effects.

### Figures

*Figure S1. Interaction effect on low-frequency (LF) gain between time segment and favorable or non-favorable ASPECTS/PC-ASPECTS in (a) $FU_{24}$-group and (b) $FU_{90}$-group. Alberta Stroke Program Early CT Score. PC-ASPECTS: Posterior circulation ASPECTS.*

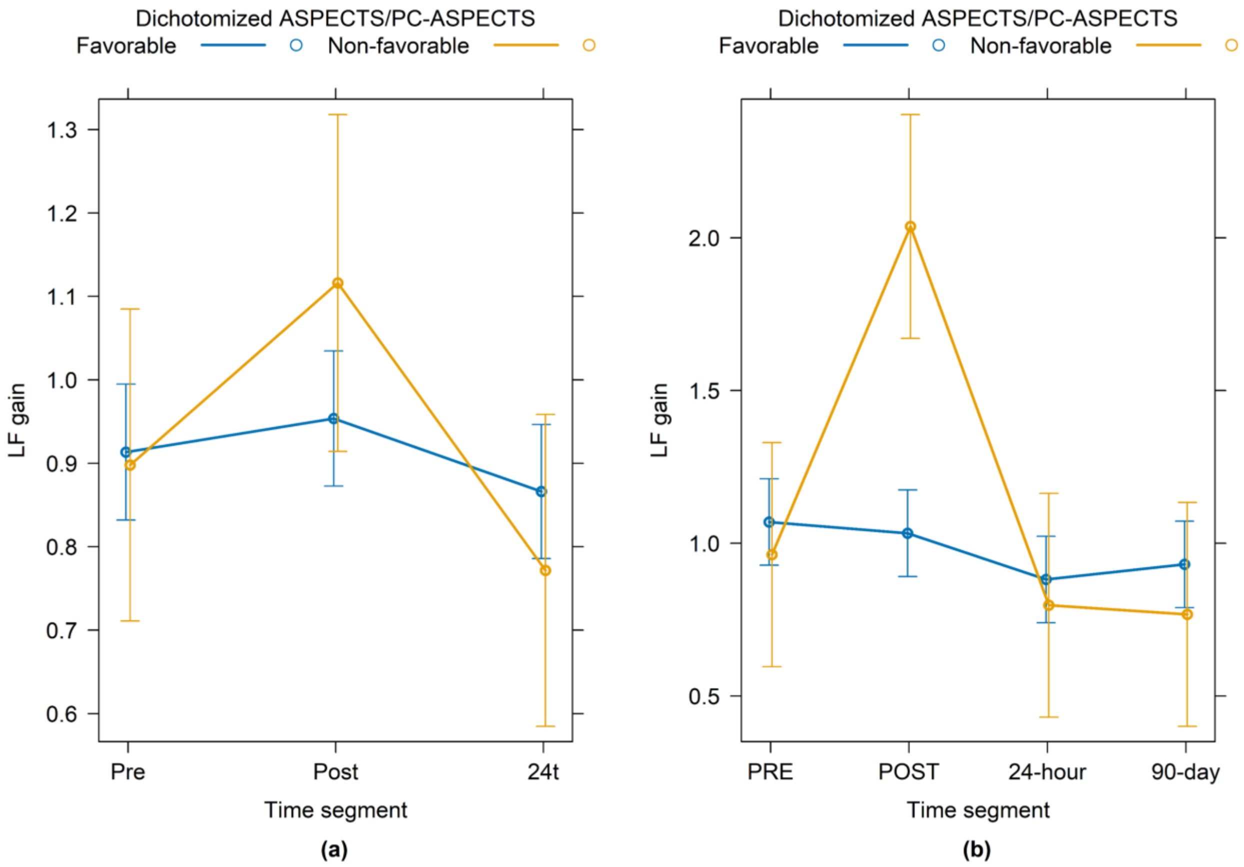

**Tables**

*Table S1. Baseline information and medical history.*

| | All enrolled (n=77) | Follow-up (n=54) | No follow-up (n=23) | P-value |
|---|---|---|---|---|
| Alcohol, weekly consumption, median units | 2 (0; 7) | 3.5 (0; 7) | 0 (0; 6) | 0.168 |
| Physical inactivity*, n (%) | 47 (61.0) | 34 (63) | 13 (56.5) | 0.618 |
| Medical history, n (%) | | | | |
| Dyslipidemia | 74 (96.1) | 53 (98.1) | 21 (91.3) | 0.211 |
| Ischemic heart disease | 12 (15.6) | 8 (14.8) | 4 (17.4) | 0.742 |
| Valvular heart disease | 9 (11.7) | 6 (11.1) | 3 (13.0) | 0.717 |
| Heart failure | 9 (11.7) | 6 (11.1) | 3 (13.0) | 0.688 |
| Pacemaker or ICD | 5 (6.5) | 4 (7.4) | 1 (4.3) | 1.000 |
| Nephropathy | 9 (11.7) | 6 (11.1) | 3 (13.0) | 1.000 |
| Venous thromboembolism | 6 (7.8) | 4 (7.4) | 1 (4.3) | 0.660 |
| Disseminated cancer | 4 (5.2) | 1 (1.9) | 3 (13.0) | 0.084 |

*BMI: Body-mass index. ICD: Implantable Cardioverter Defibrillator. *: Defined as less than 1 hour weekly. No differences between subjects with and without in-person follow-up.*

*Table S2. Index stroke treatment and outcome.*

| | All enrolled (n=77) | Follow-up (n=54) | No follow-up (n=23) | P-value |
|---|---|---|---|---|
| Procedure, n (%) | | | | |
| Aspiration only | 21 (27.3) | 15 (27.8) | 6 (26.1) | 0.540 |
| Stent-retrieving and aspiration | 52 (67.5) | 35 (64.8) | 17 (73.9) | 0.596 |
| PTA | 17 (22.1) | 15 (27.8) | 2 (8.7) | 0.078 |
| Carotid stenting | 7 (9.1) | 7 (13.0) | 0 (0) | 0.097 |
| Anesthetics, n (%) | | | | |
| Propofol | 69 (89.6) | 50 (92.6) | 19 (82.6) | 0.230 |
| Sevoflurane | 6 (7.8) | 3 (5.6) | 3 (13) | 0.356 |
| Combination of propofol and sevoflurane | 2 (2.6) | 1 (1.9) | 1 (4.3) | 0.511 |
| Vasopressor, n (%) | | | | |
| Phenylephrine | 51 (66.2) | 38 (70.4) | 13 (56.5) | 0.295 |
| Norepinephrine | 8 (10.4) | 6 (11.1) | 2 (8.7) | 1.000 |
| Phenylephrine and Norepinephrine | 18 (23.4) | 10 (18.5) | 8 (34.8) | 0.147 |
| Non-favorable ASPECTS/PC-ASPECTS before EVT (<6), n (%) | 12 (15.6) | 7 (13.0) | 5 (20.8) | 0.328 |
| Reperfusion (mTICI), n (%) | | | | 0.014 |
| 0 | 4 (5.2) | 2 (3.7) | 2 (8.7) | |
| 1 | 1 (1.3) | 0 (0) | 1 (4.3) | |
| 2a | 3 (3.9) | 1 (1.9) | 2 (8.7) | |
| 2b | 34 (44.2) | 21 (38.9) | 13 (56.5) | |
| 3 | 35 (45.5) | 30 (55.6) | 5 (21.7) | |
| Process times (minutes), median (IQR) | | | | |
| Last-known-well to imaging | 140 (79; 536) | 124 (75; 378) | 193 (91; 721) | 0.089 |
| Imaging to artery puncture | 78 (51; 108) | 68 (48; 103) | 92 (62; 110) | 0.166 |
| Artery puncture to reperfusion* | 40 (24; 69) | 36 (24; 68) | 55 (31; 72) | 0.280 |
| Complications, n (%) | | | | |
| New vascular territory embolization | 5 (6.5) | 2 (3.7) | 3 (13) | 0.154 |
| Intracranial hemorrhage | 6 (7.8) | 2 (3.7) | 4 (17.4) | 0.062 |
| Other complications† | 5 (6.5) | 3 (5.6) | 2 (8.7) | 0.632 |
| Modified Rankin Scale, median (IQR) | | | | |
| Before index stroke | 0 (0; 1) | 0 (0; 1) | 1 (0; 1.5) | 0.172 |
| Increase at 90 days | 2 (1; 3) | 1 (1; 2) | 4 (3; 5.5) | <0.001 |
| Re-hospitalized within 90 days, n (%) | 26 (33.8) | 17 (31.5) | 9 (39.1) | 0.601 |

*PTA: Percutaneous Transluminal Angioplasty. ASPECTS: Alberta Stroke Program Early CT Score. PC-ASPECTS: Posterior circulation ASPECTS. EVT: Endovascular treatment. mTICI: Modified treatment in cerebral infarction. *: In case of mTICI 0, defined as time of abandoning EVT efforts. †: Intracerebral artery dissection, vasospasm, or equipment malfunction.*

*Table S3. Median values of input and output signals, PSD in short distance channels as well as TFA gain, phase difference and coherence for all subjects at 24h follow-up.*

| Short distance channels TFA, n = 55 | | |
|---|---|---|
| Average Oxy-Hb (µM*mm) | Unaffected hemisphere | 0.01 |
| | Ischemic hemisphere | 0.02 |
| PSD ((µM*mm)$^2$ / Hz) | Unaffected hemisphere | 0.51 |
| | Ischemic hemisphere | 0.45 |
| Coherence | HF | 0.72 |
| | LF | 0.57 |
| | VLF | 0.65 |
| Gain | HF | 0.87 |
| | LF | 0.86 |
| | VLF | 0.84 |
| Phase shift (degrees) | HF | 2.78 |
| | LF | 1.04 |
| | VLF | -4.60 |

*PSD: Power spectral density. HF: High frequency spectrum (0.2-0.5 Hz). LF: Low frequency spectrum (0.07-0.2 Hz). VLF: Very-low frequency spectrum (0.02-0.07 Hz). No statistical side-to-side differences, gain different from 1 nor phase shift different from 0.*

*Table S4. Median values of input and output signals, vital parameters, PSD as well as TFA gain, phase difference and coherence for all subjects before and after reperfusion efforts and after 24 hours.*

| | | PRE (IQR) | POST (IQR) | 24-hour (IQR) |
|---|---|---|---|---|
| Examination time from last-known-well (hours) | | 4.3 (2.7; 9.7) | 5.1 (3.5; 10.4) | 27.7 (25.3; 34.9) |
| Average Oxy-Hb, unaffected hemisphere (µM*mm) | | 0.78 (-0.44; 14.73) | 0.14$^{*}$ (-2.12; 6.83) | 0.02$^{\dagger,\ddagger}$ (-2.24; 0.18) |
| Average Oxy-Hb, ischemic hemisphere (µM*mm) | | 2.36 (-0.30; 10.36) | 0.44 (-1.69; 22.25) | 0.01$^{\dagger,\ddagger}$ (-0.96; 0.08) |
| PSD, unaffected hemisphere ((µM*mm)$^2$ / Hz) | HF | 0.07 (0.03; 0.20) | 0.05 (0.03; 0.16) | 0.09 (0.03; 0.22) |
| | LF | 0.10 (0.04; 0.22) | 0.07 (0.03; 0.20) | 0.22$^{\dagger,\ddagger}$ (0.09; 0.48) |
| | VLF | 0.16 (0.08; 0.39) | 0.13$^{*}$ (0.04; 0.22) | 1.05$^{\dagger,\ddagger}$ (0.56; 1.92) |
| PSD, ischemic hemisphere ((µM*mm)$^2$ / Hz) | HF | 0.07 (0.03; 0.18) | 0.06 (0.03; 0.19) | 0.09 (0.03; 0.33) |
| | LF | 0.10 (0.05; 0.19) | 0.06 (0.03; 0.21) | 0.22$^{\dagger,\ddagger}$ (0.08; 0.53) |
| | VLF | 0.17 (0.08; 0.40) | 0.11 (0.05; 0.32) | 0.95$^{\dagger,\ddagger}$ (0.51; 1.96) |
| Coherence | HF | 0.70 (0.51; 0.89) | 0.70 (0.53; 0.88) | 0.68 (0.49; 0.80) |
| | LF | 0.54 (0.35; 0.78) | 0.55 (0.41; 0.82) | 0.53 (0.32; 0.72) |
| | VLF | 0.46 (0.27; 0.68) | 0.45 (0.27; 0.71) | 0.55 (0.39; 0.75) |
| Gain | HF | 0.92 (0.76; 1.19) | 1.00$^{*}$ (0.83; 1.37) | 0.88$^{\dagger}$ (0.76; 1.06) |
| | LF | 0.94 (0.64; 1.09) | 0.97 (0.74; 1.19) | 0.88 (0.72; 0.99) |
| | VLF | 0.81 (0.59; 1.08) | 0.87 (0.61; 1.06) | 0.83 (0.67; 0.96) |
| Phase shift (degrees) | HF | -2.02 (-6.67; 4.88) | 0.61 (-5.03; 4.81) | -1.34 (-8.57; 5.85) |
| | LF | -0.32 (-7.54; 6.53) | 2.78 (-3.79; 9.22) | -1.05 (-10.64; 10.51) |
| | VLF | -6.19 (-16.77; 3.35) | -2.79 (-7.94; 4.91) | -1.75 (-13.56; 14.31) |

| | | | | |
|---|---|---|---|---|
| Absolute phase shift (degrees) | HF | 6.11 (3.18; 11.00) | 5.03 (2.65; 10.29) | 6.66 (3.57; 12.52) |
| | LF | 6.56 (2.73; 12.10) | 7.75 (3.34; 12.47) | 10.64‡ (4.42; 19.03) |
| | VLF | 14.84 (5.84; 25.32) | 6.54* (3.73; 19.87) | 14.05† (6.92; 29.92) |
| HR (bpm, mean) | | 68.5 (SD: 13.4) | 63.7* (SD: 12.3) | 71.7†, ‡ (SD: 12.6) |
| MAP (mmHg, mean) | | 85.9 (SD: 12.1) | 82.6 (SD: 12.4) | 83.3 (SD: 13.6) |
| $SpO_2$ (%) | | 100 (98.2; 100) | 100 (98.4; 100) | 95.4†, ‡ (94; 97) |
| $ETCO_2$ (kPA) | | 4.5 (4.3; 4.76) | 4.5 (4.20; 4.76) | N/A |

*Normal distributed data specified (mean, SD). Oxy-Hb: Oxygenated hemoglobin. PSD: Power spectral density. HF: High frequency spectrum (0.2-0.5 Hz). LF: Low frequency spectrum (0.07-0.2 Hz). VLF: Very-low frequency spectrum (0.02-0.07 Hz). MAP: Mean arterial pressure. HR: Heart reate.* $SpO_2$: *Peripheral oxygen saturation.* $ETCO_2$: *End-tidal carbon dioxide. *: Significant difference between before and after reperfusion. †: Significant difference between after reperfusion and 24 hours. ‡: Significant difference between before reperfusion and 24 hours. P-value adjusted by false discovery rate.*

*Table S5. Median values of input and output signals, vital parameters, PSD as well as TFA gain, phase difference and coherence for in-person FU subjects before and after reperfusion efforts, after 24 hours and 90 days.*

| | | PRE (IQR) | POST (IQR) | 24-hour (IQR) | 90-day (IQR) |
|---|---|---|---|---|---|
| Examination time from last-known-well (hours, h or days, d) | | 4.0 h (2.5; 7.9) | 4.7 h (3.1; 8.8) | 27.4 h (25.2; 33.5) | 92.0 d (88.3; 100.8) |
| Average Oxy-Hb, unaffected hemisphere (µM*mm) | | 2.92 (-0.15; 17.48) | 1.30 (-0.23; 19.79) | 0.01 (-2.27; 0.16) | 0.02$^{*,\dagger,\ddagger}$ (-0.44; 0.56) |
| Average Oxy-Hb, ischemic hemisphere (µM*mm) | | 3.06 (-0.30; 10.36) | 0.81 (-1.35; 26.00) | 0.02 (-0.73; 0.13) | 0.02$^{*,\dagger}$ (-0.41; 0.09) |
| PSD, unaffected hemisphere ((µM*mm)$^2$/Hz) | HF | 0.06 (0.03; 0.20) | 0.05 (0.02; 0.20) | 0.11 (0.04; 0.25) | 0.06$^{\ddagger}$ (0.02; 0.22) |
| | LF | 0.10 (0.04; 0.30) | 0.10 (0.03; 0.31) | 0.25 (0.13; 0.70) | 0.19$^{\dagger,\ddagger}$ (0.07; 0.46) |
| | VLF | 0.16 (0.07; 0.43) | 0.13 (0.03; 0.26) | 1.38 (0.70; 2.00) | 0.61 (0.38; 0.91) |
| PSD, ischemic hemisphere ((µM*mm)$^2$/Hz) | HF | 0.06 (0.03; 0.25) | 0.07 (0.03; 0.28) | 0.10 (0.04; 0.37) | 0.05$^{*,\dagger,\ddagger}$ (0.02; 0.16) |
| | LF | 0.11 (0.05; 0.26) | 0.07 (0.03; 0.28) | 0.29 (0.12; 0.53) | 0.21$^{\ddagger}$ (0.06; 0.36) |
| | VLF | 0.20 (0.09; 0.42) | 0.12 (0.05; 0.36) | 1.05 (0.53; 1.96) | 0.57$^{*,\dagger,\ddagger}$ (0.40; 0.94) |
| Coherence | HF | 0.70 (0.51; 0.89) | 0.67 (0.53; 0.88) | 0.69 (0.58; 0.88) | 0.72 (0.52; 0.85) |
| | LF | 0.56 (0.36; 0.79) | 0.55 (0.42; 0.83) | 0.63 (0.39; 0.81) | 0.66 (0.53; 0.84) |
| | VLF | 0.46 (0.25; 0.70) | 0.47 (0.32; 0.71) | 0.64 (0.42; 0.78) | 0.66$^{*,\dagger}$ (0.51; 0.74) |
| Gain | HF | 0.99 (0.81; 1.21) | 1.10 (0.85; 1.39) | 0.86 (0.72; 1.05) | 0.87$^{\dagger}$ (0.79; 1.04) |
| | LF | 0.97 (0.68; 1.13) | 1.02 (0.79; 1.29) | 0.91 (0.77; 0.98) | 0.81$^{\dagger}$ (0.69; 0.98) |
| | VLF | 0.93 (0.59; 1.15) | 0.89 (0.68; 1.11) | 0.81 (0.67; 0.95) | 0.80 (0.68; 0.94) |
| Phase shift (degrees) | HF | -1.81 (-6.57; 5.64) | 0.15 (-4.93; 4.81) | -0.08 (-6.10; 6.41) | 0.92 (-4.64; 3.67) |
| | LF | -0.30 (-6.11; 6.23) | 3.91 (-3.07; 11.05) | 1.72 (-7.35; 13.89) | -3.67$^{\dagger,\ddagger}$ (-10.20; 3.73) |
| | VLF | -4.83 (-15.45; 10.34) | -2.28 (-7.49; 5.22) | 0.10 (-10.87; 14.20) | -0.49 (-11.19; 4.11) |

| | | | | | |
|---|---|---|---|---|---|
| Absolute phase shift (degrees) | HF | 6.14 (2.87; 11.11) | 4.93 (2.60; 10.29) | 6.41 (3.57; 10.04) | 4.51 (2.39; 8.83) |
| | LF | 6.23 (2.64; 11.96) | 7.84 (3.91; 12.56) | 10.51 (4.53; 15.74) | 7.66 (3.73; 11.86) |
| | VLF | 13.87 (6.14; 20.88) | 6.24 (3.80; 16.24) | 12.48 (6.56; 29.31) | 5.66*,‡ (2.70; 13.89) |
| HR (bpm, mean) | | 67.3 (SD: 13.1) | 61.8 (SD: 10) | 71.1 (SD: 12.8) | 69.7† (SD: 11.2) |
| MAP (mmHg, mean) | | 86.1 (SD: 11.8) | 80.9 (SD: 9.1) | 81.9 (SD: 13.3) | 85.3 (SD: 11.5) |
| $SpO_2$ (%) | | 100 (98; 100) | 99.8 (98; 100) | 94.5 (93.3; 97) | 96.5*,†,‡ (94.8; 98) |
| $ETCO_2$, kPA | | 4.5 () | 4.48 () | N/A | N/A |

*Normally distributed data specified (mean, SD). Oxy-Hb: Oxygenated hemoglobin. PSD: Power spectral density. HF: High frequency spectrum (0.2-0.5 Hz). LF: Low frequency spectrum (0.07-0.2 Hz). VLF: Very-low frequency spectrum (0.02-0.07 Hz). MAP: Mean arterial pressure. HR: Heart reate.* $SpO_2$: *Peripheral oxygen saturation.* $ETCO_2$: *End-tidal carbon dioxide. *: Significant difference between before reperfusion and 90 days. †: Significant difference between after reperfusion and 90 days. ‡: Significant difference between 24 hours and 90 days. P-value adjusted by false discovery rate.*

*Table S6. Mixed-effects models in $FU_{24}$-group with LF gain as outcome variable, time segment and different stroke characteristics, treatment, and outcome as fixed effect with subject as random effect.*

| Grouping | Interaction | Fixed effect<br>Groups | Fixed effect<br>Time segment<br><br>*Reference: PRE* | Model performance |
|---|---|---|---|---|
| No grouping | - | - | POST: 0.06 (CI=[-0.02; 0.15], T(147)=1.44, p = 0.152)<br><br>24-hour: -0.06 (CI=[-0.15; 0.03], T(147)=-1.35, p = 0.178) | Marginal R2 = 0.02<br>Conditional R2 = 0.34 |
| 90-day mRS | F(2, 145)=0.96, p=0.384 | -0.04 (CI=[-0.07; -0.02], T(75)=-3.2, p= 0.002)<br><br>PSD Ischemic hemisphere:<br>-0.079 (CI=[-0.156; -0.003])<br>PSD Unaffected hemisphere:<br>-0.041 (CI=[-0.099; 0.016]) | POST: 0.06 (CI=[-0.02; 0.15], T(147)=1.45, p=0.149)<br><br>24-hour: -0.06 (CI=[-0.15; 0.03], T(147)=-1.35, p=0.179) | Marginal R2 = 0.09<br>Conditional R2 = 0.34 |
| 90-day Independence (mRS 0-2) | F(2, 145)= 0.35, p=0.706 | Reference: Independence<br><br>Dependence: -0.13 (CI=[-0.24; -0.02], T(75)=-2.41 p=0.019)<br><br>PSD Ischemic hemisphere:<br>-0.328 (CI=[-0.615; -0.041])<br>PSD Unaffected hemisphere:<br>-0.148 (CI=[-0.367; 0.071]) | POST: 0.06 (CI=[-0.02; 0.15], T(147)=1.45, p=0.148)<br><br>24-hour: -0.06 (CI=[-0.15; 0.03], T(147)=-1.35, p=0.178) | Marginal R2 = 0.06<br>Conditional R2 = 0.34 |

| | | | | |
|---|---|---|---|---|
| 24-hour NIHSS recovery | F(2, 143)=0.40, p=0.673 | -0.1% (CI=[-1.2%;+1.0%], T(74)=-0.19, p=0.854) | POST: +6.8% (CI=[-3.7%; +18.3%], T(145)=1.25, p= 0.212)<br><br>24-hour: -2.3% (CI=[-11.7%; +8.2%], T(145)=-0.44, p= 0.659) | Marginal R2 = 0.01<br>Conditional R2 = 0.40 |
| NIHSS | F(2, 142)=0.25, p=0.778 | -1.0% (CI=[-1.8%; -0.1%], T(144)=-2.21, p=0.029<br><br>PSD Ischemic hemisphere:<br>-1.09e-2 (CI=[-0.015; 0.013])<br>PSD Unaffected hemisphere:<br>-1.06e-2 (CI=[-0.0190; 0.017]) | POST: +6.8% (CI=[-3.7%; +18.3%], T(145)=1.25, p= 0.215)<br><br>24-hour: -8.8% (CI=[-19.0%; +2.8%], T(145)=-1.52, p= 0.131) | Marginal R2 = 0.04<br>Conditional R2 = 0.39 |
| 90-day Mortality | F(2, 145)=1.20, p= 0.304 | Reference: Alive<br><br>Dead: -0.21 (CI=[-0.35;-0.06], T(75)=-2.91, p=0.005)<br><br>PSD Ischemic hemisphere:<br>-0.271 (CI=[-0.660; 0.117])<br>PSD Unaffected hemisphere:<br>-0.207 (CI=[-0.497; 0.082]) | POST: 0.06 (CI=[-0.02; 0.15], T(147)=1.45, p=0.149)<br><br>24-hour: -0.06 (CI=[-0.15; 0.03], T(147)=-1.37, p=0.174) | Marginal R2 = 0.08<br>Conditional R2 = 0.34 |
| Recanalization (Successful: mTICI 2b-3) | F(2, 145)=0.96, p= 0.386 | Reference: Successful<br><br>Unsuccessful: -0.15 (CI=[-0.33; 0.03], T(75)=-1.64, p=0.105)<br><br>PSD Ischemic hemisphere:<br>-0.079 (CI=[-0.564; 0.407])<br>PSD Unaffected hemisphere: | POST: 0.06 (CI=[-0.02; 0.15], T(147)=1.42, p=0.157)<br><br>24-hour: -0.06 (CI=[-0.15; 0.03], T(147)=-1.36, p=0.176) | Marginal R2 = 0.04<br>Conditional R2 = 0.34 |

| | | | | |
|---|---|---|---|---|
| | | 0.027 (CI=[-0.336; 0.390]) | | |
| IVT | F(2, 145)=0.58, p=0.554 | Reference: No IVT<br><br>IVT: 0.22 (CI=[0.12;0.32, T(75)= 4.48, p<0.001) | POST: 0.06 (CI=[-0.02; 0.15], T(147)= 1.43, p=0.154)<br><br>24-hour: -0.06 (CI=[-0.15; 0.03], T(147)=-1.36, p=0.177) | Marginal R2 = 0.14<br>Conditional R2 = 0.34 |
| ICH | F(2,145)=0.30, p=0.742 | Reference: No ICH<br><br>ICH: -0.08 (CI=[-0.29; 0.12]; T(75)=-0.79, p=0.432) | POST: 0.06 (CI=[-0.02; 0.15], T(147)= 1.44, p=0.151)<br><br>24-hour: -0.06 (CI=[-0.15; 0.03], T(147)=-1.36, p=0.177) | Marginal R2 = 0.03<br>Conditional R2 = 0.34 |
| Anesthesia | F(2, 141)=0.12, p=0.885 | Reference: Propofol<br><br>Sevoflurane: 0.01 (CI=[-0.20;0.21], T(73)=0.05, p=0.961) | POST: 0.07 (CI=[-0.02; 0.15], T(143)=1.49, p=0.139)<br><br>24-hour: -0.05 (CI=[-0.13; 0.04], T(143)=-1.05, p=0.292) | Marginal R2 = 0.02<br>Conditional R2 = 0.32 |
| Vasopressor | F(2, 110)=0.39, p=0.678 | Reference: Phenylephrine<br><br>Norepinephrine: 0.04 (CI=[-0.15; 0.24], T(57)=0.45, p=0.656) | POST: 0.07 (CI=[-0.03; 0.18], T(112)=1.42, p=0.1596)<br><br>24-hour: -0.05 (CI=[-0.16; 0.05], T(112)=-1.04, p=0.301) | Marginal R2 = 0.02<br>Conditional R2 = 0.34 |
| Stroke etiology: Large-artery atherosclerosis (LAA) or Cardio-embolic (CE) | F(2,105)=0.50, p= 0.609 | Reference: LAA<br><br>CE: -0.03 (CI=[-0.17; 0.11], T(53)=0.812, p=0.421) | POST: 0.08 (CI=[-0.02; 0.19], T(107)=1.56, p=0.116)<br><br>24-hour: -0.09 (CI=[-0.19; 0.01], T(107)=-1.72, p=0.088) | Marginal R2 = 0.04<br>Conditional R2 = 0.37 |

| | | | | |
|---|---|---|---|---|
| Age | F(2,145)=1.05, p=0.351 | -1.4e-3 (CI=[-5.6e-3; 3.5e-3], T(75)=-0.45, p=0.652) | POST: 0.06 (CI=[-0.02; 0.15], T(147)=1.44, p=0.150)<br><br>24-hour: -0.06 (CI=[-0.15; 0.03], T(147)=-1.35, p=0.178) | Marginal R2 = 0.02<br>Conditional R2 = 0.34 |
| Isolated thrombus vs complicated occlusion (ICA-Top and tandem occlusion) | F(2,133)=0.959, p=0.386 | Reference: Isolated thrombus<br><br>Complicated occlusion: 0.07 (CI=[-0.06; 0.20], T(69)=1.07, p=0.289) | POST: 0.06 (CI=[-0.03; 0.16], T(135)=1.40, p=0.162)<br><br>24-hour: -0.06 (CI=[-0.15; 0.03], T(135)=-1.31, p=0.190) | Marginal R2 = 0.03<br>Conditional R2 = 0.33 |
| ASPECTS/PC-ASPECTS before EVT (Favorable 7-10 vs non-favorable 0-6) | F(2,145)=2.16, p=0.119 | Reference: Favorable<br><br>Non-favorable: 0.01 (CI=[-0.15; 0.16], T(75)=0.11,p=0.914) | POST: 0.06 (CI=[-0.02; 0.15], T(147)=1.44, p=0.151)<br><br>24-hour: -0.06 (CI=[-0.15; 0.03], T(147)=-1.35, p=0.178) | Marginal R2 = 0.02<br>Conditional R2 = 0.34 |
| Anterior vs. posterior circulation stroke | F(2,143)=0.013, p=0.987 | Reference: Anterior<br><br>Posterior: 0.08 (CI=[-0.13; 0.29], T(74)=0.76, p=0.449) | POST: 0.07 (CI=[-0.02; 0.16], T(145)=1.51, p=0.135)<br><br>24-hour: -0.06 (CI=[-0.15; 0.06], T(145)=-1.34, p=0.182) | Marginal R2 = 0.03<br>Conditional R2 = 0.34 |
| Hypotension during EVT (MAP< 70 mmHg) | F(2,144)=0.20, p=0.817 | Reference: No hypotension<br><br>Hypotension: 0.00 (CI=[-0.11; 0.11], T(146)=-0.04, p=0.969) | POST: 0.06 (CI=[-0.02; 0.15], T(146)=1.44, p=0.153)<br><br>24-hour: -0.06 (CI=[-0.15; 0.03], T(146)= -1.28, p=0.204) | Marginal R2 = 0.02<br>Conditional R2 = 0.34 |
| Hypertension during EVT (MAP< | F(2,144)=0.26, p=0.769 | Reference: No hypertension | POST: 0.06 (CI=[-0.03; 0.14], T(146)=1.28, p=0.203) | Marginal R2 = 0.03<br>Conditional R2 = 0.34 |

| | | | | |
|---|---|---|---|---|
| 130 mmHg before recanalization or 90 mmHg after recanalization) | | Hypertension: 0.09 (CI=[-0.01; 0.19], T(146)=1.75, p=0.082)<br><br>PSD Ischemic hemisphere: 0.014 (CI=[-0.221; 0.249])<br>PSD Unaffected hemisphere: -0.031 (CI=[-0.221; 0.160]) | 24-hour: -0.06 (CI=[-0.14; 0.03], T(146)=-1.24, p=0.219) | |
| IVT and time from LKW to arrival | IVT x Time to LKW to arrival: F(1,72)= 2.54, p=0.116 | IVT: 0.21 (CI=[0.10; 0.33]; T(74)= 3.67, p<0.001)<br><br>Time to LKW to arrival: -0.00 (CI=[-0.01; 0.01]; T(73)=-0.33, p=0.741) | POST: 0.06 (CI=[-0.03; 0.15], T(147)= 1.36, p=0.175)<br><br>24-hour: -0.06 (CI=[-0.15; 0.03], T(147)=-1.36, p=0.175) | Marginal R2 = 0.13<br>Conditional R2 = 0.34 |
| IVT and ASPECTS/PC-ASPECTS before EVT | IVT x ASPECTS/PC-ASPECTS: F(1,73)=0.10, p=0.750 | IVT: 0.24 (CI=[0.13; 0.34]; T(74)= 4.51, p<0.001)<br><br>ASPECTS/PC-ASPECTS: -0.01 (CI=[-0.03; -0.01]; T(74)=-0.77, p=0.444) | POST: 0.06 (CI=[-0.02; 0.15], T(147)= 1.45, p=0.151)<br><br>24-hour: -0.06 (CI=[-0.15; 0.03], T(147)=-1.36, p=0.177) | Marginal R2 = 0.14<br>Conditional R2 = 0.35 |
| IVT and 90-day mRS | IVT x 90-day mRS: F(1,73)= 0.73, p=0.397 | IVT: 0.19 (CI=[0.09; 0.29]; T(74)= 3.67, p=0.001)<br><br>90-day mRS: -0.03 (CI=[-0.06; -0.00]; T(74)=-2.08, p=0.041) | POST: 0.06 (CI=[-0.02; 0.15], T(147)= 1.44, p=0.152)<br><br>24-hour: -0.06 (CI=[-0.15; 0.03], T(147)=-1.35, p=0.178) | Marginal R2 = 0.16<br>Conditional R2 = 0.35 |

*PSD: Power spectral density (unit: (µM*mm)$^2$ / Hz). CI: Confidence interval. mRS: Modified Rankin scale. NIHSS: National Institutes of Health Stroke Scale. mTICI: modified treatment in cerebral infarction. IVT: intravenous thrombolysis. ICH: symptomatic and non-symptomatic* *intracranial hemorrhages. ICA-top: Top of the internal carotid artery. ASPECTS: Alberta stroke program early CT score. EVT: Endovascular treatment. MAP: Mean arterial pressure. LKW: Last-known-well.*

*Table S7. Mixed-effects models in $FU_{90}$-group with LF gain as outcome variable, time segment and different stroke characteristics, treatment, and outcome as fixed effect with subject as random effect.*

| Grouping | Interaction | Fixed effect<br>Groups | Fixed effect<br>Time segment<br><br>*Reference: PRE* | Model performance |
|---|---|---|---|---|
| No grouping | - | - | POST: 0.08 (CI=[-0.04; 0.19], T(159)=1.36, p = 0.184)<br><br>24-hour: -0.09 (CI=[-0.20; 0.02], T(159)=-1.65, p=0.100)<br><br>90-day: -0.10 (CI=[-0.21; 0.01], T(159)=-1.79, p=0.076) | Marginal R2 = 0.05<br>Conditional R2 = 0.12 |
| 90-day mRS | F(3,150)=0.16, p=0.921 | -0.03 (CI=[-0.07; 0.00], T(52)=-1.85, p=0.070) | POST: 0.08 (CI=[-0.04; 0.19], T(153)=1.35, p=0.180)<br><br>24-hour: -0.09 (CI=[-0.20; 0.02], T(153)=-1.65, p=0.101)<br><br>90-day: -0.10 (CI=[-0.21; 0.01], T(153)=-1.77, p=0.078) | Marginal R2 = 0.07<br>Conditional R2 = 0.21 |
| 90-day Independence (mRS 0-2) | F(3,150)=0.23, p=0.874 | Reference: Independence<br><br>Dependence: -0.10 (CI=[-0.20;-0.01], T(52)=-1.87, p=0.067 | POST: 0.08 (CI=[-0.04; 0.19], T(153)=1.34, p=0.180)<br><br>24-hour: -0.09 (CI=[-0.20; 0.02], T(153)=-1.65, p=0.101) | Marginal R2 = 0.07<br>Conditional R2 = 0.21 |

| | | | | |
|---|---|---|---|---|
| | | | 90-day: -0.10 (CI=[-0.21; 0.01], T(153)=-1.77, p=0.078) | |
| 90-day NIHSS recovery | F(3,150)=0.08, p=0.973 | +0.1% (CI=[-0.9%; +1.2%], T(52)=0.28, p=0.783) | POST: +5.6% (CI=[-6.5%; +19.3%], T(153)=0.89, p=0.374)<br><br>24-hour: -6.2% (CI=[-16.8%; +5.7%], T(153)=-1.06, p=0.291)<br><br>90-day: -7.2% (CI=[-17.7%; +5.8%], T(153)=-1.21, p=0.227) | Marginal R2 = 0.02 Conditional R2 = 0.21 |
| NIHSS | F(3,149)=0.15, p=0.932 | -0.5% (CI=[-1.4%;+0.5%], T(152)=-0.96, p=0.337) | POST: +5.6% (CI=[-6.5%;+19.3%], T(152)=0.89, p=0.377)<br><br>24-hour: -9.8% (CI=[-21.8%; +4.2%], T(152)=-1.41, p=0.160)<br><br>90-day: -11.9% (CI=[-25.1%; +3.6%], T(152)=-1.54, p=0.125) | Marginal R2 = 0.03 Conditional R2 = 0.20 |
| Recanalization (Successful: mTICI 2b-3) | F(3,150)=0.77, p=0.515 | Reference: Successful<br><br>Unsuccessful: -0.17 (CI=[-0.06; 0.40], T(52)=1.47, p=0.148) | POST: 0.08 (CI=[-0.04; 0.19], T(153)=1.30, p=0.195)<br><br>24-hour: -0.09 (CI=[-0.20;0.02], T(153)=-1.66, p=0.099)<br><br>90-day: -0.10 (CI=[-0.21; 0.01], T(153)=-1.79, p=0.075) | Marginal R2 = 0.06 Conditional R2 = 0.21 |
| IVT | F(3,150)= 0.86, p=0.464 | Reference: No IVT | POST: 0.07 (CI=[-0.04; 0.19], T(153)= 1.33, p=0.187) | Marginal R2 = 0.11 Conditional R2 = 0.21 |

| | | | | |
|---|---|---|---|---|
| | | IVT: 0.16 (CI=[0.07;0.26, T(52)= 2.63, p=0.011) | 24-hour: -0.09 (CI=[-0.20;0.02], T(153)=-1.65, p=0.100)<br><br>90-day: -0.10 (CI=[-0.21; 0.01], T(153)= -1.76, p=0.078) | |
| Anesthesia | F(3,147)=0.542, p=0.654 | Reference: Propofol<br><br>Sevoflurane: -0.02 (CI=[-0.25;0.22], T(51)=-0.13, p=0.894) | POST: 0.08 (CI=[-0.04; 0.19], T(153)=1.34, p=0.184)<br><br>24-hour: -0.08 (CI=[-0.19;0.03], T(153)=-1.40, p=0.165)<br><br>90-day: -0.08 (CI=[-0.19; 0.02], T(153)=-1.50, p=0.136) | Marginal R2 = 0.04<br>Conditional R2 = 0.20 |
| Vasopressor | F(3, 121)=0.59, p=0.623 | Reference: Phenylephrine<br><br>Norepinephrine: -0.02 (CI=[-0.20; 0.15], T(42)=-0.28, p=0.779) | POST: 0.08 (CI=[-0.04; 0.21], T(124)=1.33, p=0.185)<br><br>24-hour: -0.09 (CI=[-0.22; 0.03], T(124)=-1.53, p=0.128)<br><br>90-day: -0.11 (CI=[-0.23; 0.01], T(124)=-1.78, p=0.078) | Marginal R2 = 0.06<br>Conditional R2 = 0.20 |
| Stroke etiology: Large-artery atherosclerosis (LAA) or Cardio-embolic (CE) | F(3,109)=0.17, p=0.917 | Reference: LAA<br><br>CE: -0.04 (CI=[-0.18;0.10], T(37)=-0.62, p=0.538) | POST: 0.08 (CI=]-0.05; 0.21], T(112)=1.29, p=0.200)<br><br>24-hour: -0.14 (CI=[-0.27;-0.01], T(112)=-2.16, p=0.033)<br><br>90-day: -0.14 (CI=[-0.27; -0.01], T(112)=-2.18, p=0.031) | Marginal R2 = 0.08<br>Conditional R2 = 0.29 |

| | | | | |
|---|---|---|---|---|
| Age | F(3,150)=0.64, p=0.587 | 0.00 (CI=[-0.01; 0.00], T(52)=-0.72, p=0.474) | POST: 0.08 (CI=[-0.04; 0.19], T(153)=1.35, p=0.179)<br><br>24-hour: -0.09 (CI=[-0.20;0.02], T(153)=-1.65, p=0.101)<br><br>90-day: -0.10 (CI=[-0.21; 0.01], T(153)=-1.79, p=0.075) | Marginal R2 = 0.02<br>Conditional R2 = 0.21 |
| Isolated thrombus vs complicated occlusion (ICA-Top and tandem occlusion) | F(3,147)=1.68, p=0.173 | Reference: Isolated thrombus<br><br>Complicated occlusion:<br>-11.0% (CI=[-4.0%; +28.3%], T(49)=-1.45, p=0.155) | POST: +8.6% (CI=[-6.5%; +26.2%], T(150)=1.09, p=0.277)<br><br>24-hour: -9.8% (CI=[-22.4%; +4.7%], T(150)=-1.37, p=0.174)<br><br>90-day: -7.8% (CI=[-20.6%; +7.1%], T(150)=-1.08 p=0.283) | Marginal R2 = 0.04<br>Conditional R2 = 0.15 |
| ASPECTS/PC-ASPECTS before EVT (Favorable 7-10 vs non-favorable 0-6) | F(3,150)=2.95, p=0.035 | Reference: Favorable<br><br>Non-favorable: -0.02 (CI=[-0.10; 0.06], T(52)=-0.55, p=0.585) | POST: 0.04 (CI=[-0.08; 0.16], T(150)=0.20, p=0.844)<br><br>24-hour: -0.08 (CI=[-0.20; 0.03], T(150)=4.17, p<0.001)<br><br>90-day: -0.09 (CI=[-0.20; 0.03], T(150)=-2.26, p=0.025) | Marginal R2 = 0.08<br>Conditional R2 = 0.24 |

| | | | | |
|---|---|---|---|---|
| Anterior vs. posterior circulation stroke | F(3,149)=1.01, p=0.391 | Reference: Anterior<br><br>Posterior: 0.02 (CI=[-0.12; 0.16], T(152)=0.30, p=0.765) | POST: 0.08 (CI=[-0.04; 0.19], T(152)=1.34, p=0.181)<br><br>24-hour: -0.09 (CI=[-0.20; 0.02], T(152)=-1.64, p=0.103)<br><br>90-day: -0.10 (CI=[-0.21;0.01], T(152)=-1.78, p=0.077) | Marginal R2 = 0.05<br>Conditional R2 = 0.20 |
| Hypotension during EVT (MAP< 70 mmHg) | Not evaluable | Reference: No hypotension<br><br>Hypoperfusion: -0.03 (CI=[-0.17; 0.10], T(152)=-0.50, p= 0.621) | POST: 0.07 (CI=[-0.04; 0.19], T(149)=1.31, p=0.192)<br><br>24-hour: -0.09 (CI=[-0.20;0.02], T(149)=-1.63, p=0.105)<br><br>90-day: -0.09 (CI=-[0.21; 0.04], T(149)=-1.37, p=0.174) | Marginal R2 = 0.05<br>Conditional R2 = 0.21 |
| Hypertension during EVT (MAP< 130 mmHg before recanalization or 90 mmHg after recanalization) | F(3,149)=1.43, p=0.235 | Reference: No hypertension<br><br>Hypertension: 0.09 (CI=[-0.02; 0.20], T(152)=1.65, p=0.102) | POST: 0.08 (CI=[-0.03; 0.20], T(152)=1.49, p=0.138)<br><br>24-hour: -0.08 (CI=[-0.19;0.03], T(152)=-1.51, p=0.134)<br><br>90-day: -0.09 (CI=[-0.20; 0.02], T(152)=-1.61, p=0.110) | Marginal R2 = 0.06<br>Conditional R2 = 0.21 |

*Power spectral density (unit: (µM*mm)$^2$ / Hz). CI: Confidence interval. mRS: Modified Rankin scale. NIHSS: National Institutes of Health Stroke Scale. mTICI: modified treatment in cerebral infarction. IVT: intravenous thrombolysis. ICA-top: Top of the internal carotid artery. ASPECTS: Alberta stroke program early CT score. EVT: Endovascular treatment. MAP: Mean arterial pressure.*

*Table S8. Mixed-effects models in $FU_{24}$-group with LF absolute phase shift as outcome variable, time segment and different stroke characteristics, treatment, and outcome as fixed effect with subject as random effect.*

| Grouping | Interaction | Fixed effect<br>Groups | Fixed effect<br>Time segment<br><br>*Reference: PRE* | Model performance |
|---|---|---|---|---|
| No grouping | - | - | POST: + 8.2% (CI=[-24.0%; +54.1%], T(152)=0.44, p = 0.661)<br><br>24-hour: +56.0% (CI=[+9.6%; +122.2%], T(152)=2.49, p=0.014) | Marginal R2 = 0.03<br>Conditional R2 = 0.16 |
| 90-day mRS | F(2,150)=1.09, p=0.339 | 0.0% (CI=[-9.0%; +9.9%], T(75)=0.00, p=0.997) | POST: +8.2% (CI=[-24.0%; +54.1%, T(152)=0.44, p=0.661)<br><br>24-hour: +56.0 (CI=+9.6%; +122.2%], T(152)=2.49, p=0.014) | Marginal R2 = 0.03<br>Conditional R2 = 0.16 |
| 90-day Independence (mRS 0-2) | F(2,150)=1.24, p=0.293 | Reference: Independence<br><br>Dependence: -3.2% (CI=[-47.3%; +27.7%], T(75)=-0.18, p=0.860) | POST: +8.2% (CI=[-24.0%; +54.1%], T(152)=0.44, p=0.661)<br><br>24-hour: +56.0 (CI=[+9.6%; +122.2%], T(152)=2.49, p=0.014) | Marginal R2 = 0.03<br>Conditional R2 = 0.17 |

| | | | | |
|---|---|---|---|---|
| 24-hour NIHSS recovery | F(2,148)=0.62, p=0.541 | +0.5% (CI=[-3.6%;+4.8%], T(74)=0.24, p=0.809) | POST: +7.5% (CI=[-36.8%; +82.8%], T(148)=0.27, p=0.788)<br><br>24-hour: +89.5% (CI=[+11.4%; +222.2%], T(148)=2.38, p=0.019) | Marginal R2 = 0.03<br>Conditional R2 = 0.16 |
| NIHSS | F(2,147)=3.96, p=0.021 | +4.6% (CI=[0.4%; +8.9%], T(147)=2.16, p=0.032) | POST: +257.1% (CI=[+42.2%; +796.8%], T(147)=2.73, p=0.007)<br><br>24-hour: +181.7% (CI=[+28.6%; +516.8%], T(147)=2.61, p=0.010) | Marginal R2 = 0.06<br>Conditional R2 = 0.20 |
| 90-day Mortality | F(2,150)=0.11, p=0.894 | Reference: Alive<br><br>Dead: +24.2% (CI=[-22.5; +99.0%], T(75)=0.92, p=0.363) | POST: +8.2% (CI=[-24.0%; +54.1%], T(152)=0.44, p=0.661)<br><br>24-hour: +56.0% (CI=[+9.6%; +122.2%], T(152)=2.49, p=0.014) | Marginal R2 = 0.03<br>Conditional R2 = 0.16 |
| Recanalization (Successful: mTICI 2b-3) | F(2,150)=0.80, p=0.451 | Reference: Successful<br><br>Unsuccessful: +32.6% (CI=[-25.6%; +136.3%], T(75)=0.97, p=0.335) | POST: +8.2% (CI=[-24.0%; +54.1%], T(152)=0.44, p=0.661)<br><br>24-hour: +56.0% (CI=]+9.6%; +122.2%], T(152)=2.49, p=0.014) | Marginal R2 = 0.03<br>Conditional R2 = 0.16 |

| | | | | |
|---|---|---|---|---|
| IVT | F(2,150)= 0.11, p=0.900 | Reference: No IVT<br><br>IVT: -5.5% (CI=[-33.8%; +35.0%]; T(75)=-0.32, p=0.753) | POST: +8.2% (CI=[-24.0%; +54.1%], T(152)=0.44, p=0.661)<br><br>24-hour: +56.0% (CI=]+9.6%; +122.2%], T(152)=2.49, p=0.014) | Marginal R2 = 0.03<br>Conditional R2 = 0.16 |
| ICH | F(2,150)=0.32, p=0.729 | Reference: No ICH<br><br>ICH: +16.1% (CI=[-40.1%; +125.0%]; T(75)=0.45, p=0.654) | POST: +8.2% (CI=[-24.0%; +54.1%], T(152)=0.44, p=0.661)<br><br>24-hour: +56.0% (CI=]+9.6%; +122.2%], T(152)=2.49, p=0.014) | Marginal R2 = 0.03<br>Conditional R2 = 0.16 |
| Anesthesia | F(2,146)=1.28, p=0.281 | Reference: Propofol<br><br>Sevoflurane: -46.0% (CI=[-71.9%; +3.9%], T(73)=-1.88, p=0.064) | POST: +7.0% (CI=[-25.2%; +53.2%], T(148)=0.37, p=0.709)<br>24-hour: +61.4% (CI=[+12.8%; +131.0%], T(148)=2.64, p=0.009) | Marginal R2 = 0.05<br>Conditional R2 = 0.17 |
| Vasopressor | F(2, 114)=1.71, p=0.186 | Reference: Phenylephrine<br><br>Norepinephrine: -3.8% (CI=[-47.8%; +77.0%], T(57)=-0.13, p=0.899) | POST: +21.5% (CI=[-21.5%; +88.1%], T(116)=0.88, p=0.378)<br><br>24-hour: +61.8% (CI=[+4.6%; +150.4%], T(116)=2.18, p=0.031) | Marginal R2 = 0.02<br>Conditional R2 = 0.12 |

| | | | | |
|---|---|---|---|---|
| Stroke etiology: Large-artery atherosclerosis (LAA) or Cardio-embolic (CE) | F(2,106)=1.36, p=0.262 | Reference: LAA<br><br>CE: -16.6% (CI=[-43.6%; +23.2%], T(53)=-0.93, p=0.355) | POST: +38.3% (CI=[-9.8%; +112.1%], T(108)=1.50, p=0.136)<br><br>24-hour: +62.0% (CI=[+5.6%; +148.4%], T(108)=2.23, p=0.028) | Marginal R2 = 0.03<br>Conditional R2 = 0.10 |
| Age | F(2,150)=0.04, p=0.965 | -0.3% (CI=[-1.8%; +1.1%], T(75)=-0.45, p=0.653) | POST: +8.2% (CI=[-24.0%; +54.1%], T(152)=0.44, p=0.661)<br><br>24-hour: +56.0% (CI=[+9.6%; +122.2], T(152)=2.49, p=0.014) | Marginal R2 = 0.03<br>Conditional R2 = 0.16 |
| Isolated thrombus vs complicated occlusion (ICA-Top and tandem occlusion) | F(2,138)=0.027, p=0.973 | Reference: Isolated thrombus<br><br>Complicated occlusion: -22.8% (CI=[-48.9%; +16.7%], T(69)=-1.25, p=0.216) | POST: +7.8% (CI=[-25.9%; +56.9%], T(140)=0.40, p=0.693)<br><br>24-hour: +51.5% (CI=[+4.1%; +120.5%], T(140)=2.19, p=0.030) | Marginal R2 = 0.03<br>Conditional R2 = 0.16 |
| ASPECTS/PC-ASPECTS before EVT (Favorable 7-10 vs non-favorable 0-6) | F(2,150)=0.40, p=0.669 | Reference: Favorable<br><br>Non-favorable: +21.5% (CI=[-25.4%; +97.8%], T(75)=0.80, p=0.428) | POST: +8.2% (CI=[-24.0%; +54.1%], T(152)=0.44, p=0.661)<br><br>24-hour: +56.0% (CI=[+9.6%; +122.2%], T(152)=2.49, p=0.014) | Marginal R2 = 0.03<br>Conditional R2 = 0.16 |

| | | | | |
|---|---|---|---|---|
| Anterior vs. posterior circulation stroke | F(2,148)=0.21, p=0.808 | Reference: Anterior<br><br>Posterior: +27.8% (CI=[-34.2; +148.4%], T(74)=0.74, p=0.464) | POST: +3.8% (CI=[-27.1%; +47.8%], T(150)=0.21, p=0.833)<br><br>24-hour: +51.4% (CI=[+6.3%; +115.5%], T(150)=2.32, p=0.022) | Marginal R2 = 0.03<br>Conditional R2 = 0.17 |
| Hypotension during EVT (MAP< 70 mmHg) | F(2,149)=0.32, p=0.725 | Reference: No hypotension<br><br>Hypotension: +14.2% (CI=[-25.2%; +74.5%], T(151)=0.62, p=0.536) | POST: +7.6% (CI=[-24.5%; +53.3%], T(151)=0.41, p=0.682)<br><br>24-hour: +51.3% (CI=[+4.8%; +118.4%], T(151)=2.23, p=0.027) | Marginal R2 = 0.03<br>Conditional R2 = 0.16 |
| Hypertension during EVT (MAP< 130 mmHg before recanalization OR 90 mmHg after recanalization) | F(2,149)=1.09, p=0.339 | Reference: No hypertension<br><br>Hyperpertension: +7.9% (CI=[-26.6%; +58.6%], T(151)=0.39, p=0.697) | POST: +7.4% (CI=[-24.7%; +53.3%], T(151)=0.40, p=0.697)<br><br>24-hour: +56.7% (CI=[+9.9%; +123.3%], T(151)=2.50, p=0.013) | Marginal R2 = 0.03<br>Conditional R2 = 0.16 |

*PSD: Power spectral density (unit: (μM*mm)$^2$ / Hz). CI: Confidence interval. mRS: Modified Rankin scale. NIHSS: National Institutes of Health Stroke Scale. mTICI: modified treatment in cerebral infarction. IVT: intravenous thrombolysis. ICH: symptomatic and non-symptomatic intracranial hemorrhages. ICA-top: Top of the internal carotid artery. ASPECTS: Alberta stroke program early CT score. EVT: Endovascular treatment. MAP: Mean arterial pressure.*

*Table S9. Mixed-effects models in $FU_{90}$-group with LF absolute phase shift as outcome variable, time segment and different stroke characteristics, treatment, and outcome as fixed effect with subject as random effect.*

| Grouping | Interaction | Fixed effect<br>Grupper | Fixed effect<br>Time segment<br><br>*Reference: PRE* | Model performance |
|---|---|---|---|---|
| No grouping | - | - | POST: +28.0% (CI=[-14.8%; +92.3%], T(159)=1.20, p=0.233)<br><br>24-hour: +66.1% (CI=[+10.5%; +149.5%], T(159)=2.46, p=0.015)<br><br>90-day: +27.3% (CI=[-15.3%; +91.3%], T(159)=1.17, p=0.243) | Marginal R2 = 0.02<br>Conditional R2 = 0.17 |
| 90-day mRS | F(3,156)=0.82, p=0.485 | -8.6% (CI=[-20.1%; +4.5%], T(52)=-1.35, p=0.183) | POST: +28.0% (CI=[-14.8%; +92.3%], T(159)=1.20, p=0.233)<br><br>24-hour: +66.1% (CI=[+10.5%; +149.5%], T(159)=2.46, p=0.015)<br><br>90-day: +27.3% (CI=[-15.3%; +91.3%], T(159)=1.18, p=0.243) | Marginal R2 = 0.03<br>Conditional R2 = 0.17 |
| 90-day Independence (mRS 0-2) | F(3,156)=1.02, p=0.385 | Reference: Independence | POST: +28.0% (CI=[-14.8%; +92.3%], T(159)=1.20, p=0.233) | Marginal R2 = 0.03<br>Conditional R2 = 0.17 |

| | | | | |
|---|---|---|---|---|
| | | Dependence: -9.7% (CI=[-38.9%; +33.4%], T(52)=-0.53, p=0.601) | 24-hour: +66.1% (CI=[+10.5%; +149.5%], T(159)=2.46, p=0.015)<br><br>90-day: +27.3% (CI=[-15.3%; +91.3%], T(159)=1.18, p=0.243) | |
| 90-day NIHSS recovery | F(3,156)=1.51, p=0.215 | -0.5% (CI=[-3.7%; +2.8%], T(52)=-0.31, p=0.756) | POST: +28.0% (CI=[-14.8%; +92.3%], T(159)=1.20, p=0.233)<br><br>24-hour: +66.1% (CI=[+10.5%; +149.5%], T(159)=2.46, p=0.015)<br><br>90-day: +27.3% (CI=[-15.3%; +91.3%], T(159)=1.17, p=0.243) | Marginal R2 = 0.03<br>Conditional R2 = 0.17 |
| NIHSS | F(3,155)=3.82, p=0.011 | +3.5% (CI=[-1.4%; +8.6%], T(155)=1.42, p=0.158) | POST: +414.1% (CI=[+84.6%; +1331.7%], T(155)=3.16, p=0.002)<br><br>24-hour: +115.6% (CI=[-10.0%; +416.2%], T(155)=1.74, p=0.084)<br><br>90-day: +95.8% (CI=[-15.2%; +351.9%], T(155)=1.59, p=0.115) | Marginal R2 = 0.07<br>Conditional R2 = 0.22 |
| Recanalization (Successful: mTICI 2b-3) | F(3,156)=1.63, p=0.186 | Reference: Successful | POST: +28.0% (CI=[-14.8%; +92.3%], T(159)=1.20, p=0.233) | Marginal R2 = 0.02<br>Conditional R2 = 0.17 |

| | | | | |
|---|---|---|---|---|
| | | Unsuccessful: +15.5% (CI=[-95.2%; +63.4%], T(52)=0.40, p=0.688) | 24-hour: +66.1% (CI=[+10.5%; +149.5%], T(159)=2.46, p=0.015)<br><br>90-day: +27.3% (CI=[-15.3%; +91.3%], T(159)=1.17, p=0.243) | |
| Intravenous thrombolysis (IVT) | F(3,156)=0.16, p=0.924 | Reference: No IVT<br><br>IVT: +10.1% (CI=[-25.0%; +61.6%]; T(52)=0.50, p=0.616) | POST: +28.0% (CI=[-14.8%; +92.3%], T(159)=1.20, p=0.233)<br><br>24-hour: +66.1% (CI=[+10.5%; +149.5%], T(159)=2.46, p=0.015)<br><br>90-day: +27.3% (CI=[-15.3%; +91.3%], T(159)=1.17, p=0.243) | Marginal R2 = 0.02<br>Conditional R2 = 0.17 |
| Anesthesia | F(3,153)=1.16, p=0.327 | Reference: Propofol<br><br>Sevoflurane: -39.1% (CI=[-73.6%; +40.2%], T(51)=-1.19, p=0.238) | POST: +26.5% (CI=[-16.4%; +91.6%], T(156)=1.12, p=0.264)<br><br>24-hour: +66.3% (CI=[+9.9%; +151.8%], T(156)=2.42, p=0.017)<br><br>90-day: +27.5% (CI=[-15.8%; +93.0%], T(156)=1.16, p=0.249) | Marginal R2 = 0.03<br>Conditional R2 = 0.17 |
| Vasopressor | F(3, 126)=0.31, p=0.820 | Reference: Phenylephrine<br><br>Norepinephrine: +23.4% (CI=[-34.6%; +133.0%], T(42)=0.67, p=0.508) | POST: +45.9% (CI=[-34.6%; +133.0%], T(129)=1.57, p=0.118) | Marginal R2 = 0.03<br>Conditional R2 = 0.16 |

| | | | | |
|---|---|---|---|---|
| | | | 24-hour: +75.8% (CI=[+9.3%; +182.8%], T(129)=2.35, p=0.020)<br><br>90-day: +43.0% (CI=[-11.1%; +130.0%], T(129)=1.49, p=0.139) | |
| Stroke etiology: Large-artery atherosclerosis (LAA) vs Cardio-embolic (CE) | F(3,111)= 1.08, p=0.363 | Reference: LAA<br><br>CE: -22.7% (CI=[-51.7; +23.7%], T(37)=-1.11, p=0.275) | POST: +67.6% (CI=[+4.1%; +169.8%], T(114)=2.15, p=0.034)<br><br>24-hour: +82.8% (CI=[+13.6%; +194.2%], T(114)=2.51, p=0.014)<br><br>90-day: +50.2% (CI=[-6.7%; +141.8%], T(114)=1.69, p=0.093) | Marginal R2 = 0.05<br>Conditional R2 = 0.22 |
| Age | F(3,156)=0.23, p=0.873 | -0.1% (CI=[-1.6%;+1.5%], T(52)=-0.08, p=0.935) | POST: +28.0% (CI=[-14.8%; +92.3%], T(159)=1.20, p=0.233)<br><br>24-hour: +66.1% (CI=[+10.5%; +149.5%], T(159)=2.46, p=0.015)<br><br>90-day: +27.3% (CI=[-15.3%; +91.3%], T(159)=1.17, p=0.243) | Marginal R2 = 0.02<br>Conditional R2 = 0.17 |
| Isolated thrombus vs complicated | F(3,147)=0.32, p=0.808 | Reference: Isolated thrombus | POST: +21.5% (CI=[-20.2%; +85.0%], T(150)=0.91, p=0.362) | Marginal R2 = 0.03<br>Conditional R2 = 0.18 |

| | | | | |
|---|---|---|---|---|
| occlusion (ICA-Top and tandem occlusion) | | Complicated occlusion: -27.8% (CI=[-53.1%; +12.4%], T(49)=-1.47, p=0.147) | 24-hour: +55.9% (CI=[+2.4%; +137.4%], T(150)=2.09, p=0.039)<br><br>90-day: +23.2% (CI=[-19.1%; +87.6%], T(150)= 0.98, p=0.329) | |
| ASPECTS/PC-ASPECTS before EVT (Favorable 7-10 vs non-favorable 0-6) | F(3,156)=0.97, p=0.408 | Reference: Favorable<br><br>Non-favorable: +28.6% (CI=[-27.1%; +126.9%], T(52)=0.89, p=0.378) | POST: +28.0% (CI=[-14.8%; +92.3%], T(159)=1.20, p=0.233)<br><br>24-hour: +66.1% (CI=[+10.5%; +149.5%], T(159)=2.46, p=0.015)<br><br>90-day: +27.3% (CI=[-15.3%; +91.3%], T(159)=1.17, p=0.243) | Marginal R2 = 0.03<br>Conditional R2 = 0.17 |
| Anterior vs. posterior circulation stroke | F(3,155)= 0.22, p=0.884 | Reference: Anterior<br><br>Posterior: -29.1% (CI=[-57.4%; +18.1%], T(158)=-1.33, p=0.186) | POST: +26.4% (CI=[-16.0%; +90.1%], T(158)=1.13, p=0.259)<br><br>24-hour: +65.0% (CI=[+9.8%; +148.1%], T(158)=2.43, p=0.016)<br><br>90-day: +27.3% (CI=[-15.3%; +91.4%], T(158)=1.17, p=0.244) | Marginal R2 = 0.03<br>Conditional R2 = 0.17 |
| Hypotension during EVT (MAP< 70 mmHg) | Not evaluable | Reference: No hypotension | POST: +31.2% (CI=[-12.8%; +97.4%], T(158)=1.31, p=0.191) | Marginal R2 = 0.03<br>Conditional R2 = 0.17 |

| | | | | |
|---|---|---|---|---|
| | | Hypotension: +39.7% (CI=[-13.5%; +125.7%], T(158)= 1.38, p=.170) | 24-hour: +66.1% (CI=[+10.6%; +149.4%], T(158)=2.46, p=0.015)<br><br>90-day: +12.5% (CI=[-15.3%; +91.3%], T(158)=0.52, p=0.601) | |
| Hypertension during EVT (MAP< 130 mmHg before recanalization OR 90 mmHg after recanalization) | F(3,155)=0.56, p=0.639 | Reference: No hypertension<br><br>Hypertension: -4.9% (CI=[-36.6%; +42.7%], T(158)=-0.24, p=0.807) | POST: +27.5% (CI=[-15.3%; +92.0%], T(158)=1.17, p=0.242)<br><br>24-hour: +65.4% (CI=[+9.9%; +149.1%], T(158)=2.42, p=0.016)<br><br>90-day: +26.9% (CI=[-15.8%; +91.0%], T(158)=1.15, p=0.253) | Marginal R2 = 0.02<br>Conditional R2 = 0.17 |

*PSD: Power spectral density (unit: (µM*mm)$^2$ / Hz). CI: Confidence interval. mRS: Modified Rankin scale. NIHSS: National Institutes of Health Stroke Scale. mTICI: modified treatment in cerebral infarction. ICA-top: Top of the internal carotid artery. ASPECTS: Alberta stroke program early CT score. EVT: Endovascular treatment. MAP: Mean arterial pressure.*

*Table S10. Acute (logistic regression) model predicting 90-day independency (mRS 0-2).*

| Predictor | Estimate | SE | Z-value | P-value |
|---|---|---|---|---|
| LF gain | 2.601 | 1.234 | 2.107 | 0.035 |
| Age | -0.086 | 0.025 | -3.430 | 0.001 |
| Recanalization (successful) | 0.738 | 1.031 | 0.716 | 0.474 |
| ASPECTS/PC-ASPECTS (non-favorable) | -1.176 | 0.825 | -1.426 | 0.154 |
| Intercept: 90-day independency (mRS 0-2) | 3.120 | 2.217 | 1.407 | 0.159 |
| AIC | 93.127 | | | |
| Residual deviance | 83.127 | | | |

*AIC: Akaike information criterion.*

*Table S11. 24-hour (logistic regression) model predicting 90-day independency (mRS 0-2).*

| Predictor | Estimate | SE | Z-value | P-value |
|---|---|---|---|---|
| LF gain | 2.294 | 1.316 | 1.744 | 0.081 |
| Age | -0.087 | 0.029 | -3.047 | 0.002 |
| 24-hour NIHSS: Moderate | -2.235 | 0.688 | -3.247 | 0.001 |
| 24-hour NIHSS: Severe | -3.054 | 0.934 | -3.269 | 0.001 |
| Intercept: 90-day independency (mRS 0-2) | 5.488 | 2.414 | | |
| AIC | 79.958 | | | |
| Residual deviance | 65.958 | | | |

*AIC: Akaike information criterion.*

*Table S12. Acute (ordinal logistic regression) model predicting 90-day categorized NIHSS.*

| Predictor | Estimate | SE | T-value | P-value |
|---|---|---|---|---|
| LF gain | 2.264 | 0.947 | 2.390 | 0.017 |
| Age | -0.025 | 0.018 | -1.397 | 0.162 |
| Recanalization (successful) | 1.368 | 0.727 | 1.881 | 0.060 |
| ASPECTS/PC-ASPECTS (non-favorable) | -1.542 | 0.635 | -2.429 | 0.015 |
| Intercepts: Categorized 90-day NIHSS | Estimate | SE | | |
| Fatal \| Severe | -0.478 | 1.695 | | |
| Severe \| Moderate | 0.366 | 1.701 | | |
| Moderate \| Mild | 1.164 | 1.719 | | |
| Mild \| Near-remission | 2.003 | 1.732 | | |
| AIC | 231.686 | | | |
| Residual deviance | 215.686 | | | |

*AIC: Akaike information criterion.*

*Table S13. 24-hour (ordinal logistic regression) model predicting 90-day categorized NIHSS.*

| Predictors (index) | Estimate | SE | T-value | P-value |
|---|---|---|---|---|
| LF gain | 1.578 | 1.048 | 1.506 | 0.132 |
| Age | -0.016 | 0.019 | -0.874 | 0.382 |
| 24-hour NIHSS: Moderate | -2.682 | 0.591 | -4.536 | <0.001 |
| 24-hour NIHSS: Severe | -4.236 | 0.762 | -5.562 | <0.001 |
| Intercepts: Categorized 90-day NIHSS | | | | |
| Fatal \| Severe | -4.000 | 1.919 | | |
| Severe \| Moderate | -2.875 | 1.899 | | |
| Moderate \| Mild | -1.702 | 1.900 | | |
| Mild \| Near-remission | -0.407 | 1.896 | | |
| AIC | 213.285 | | | |
| Residual deviance | 193.285 | | | |

*AIC: Akaike information criterion.*

*Table S14. Acute (logistic regression) model predicting 90-day mortality.*

| Predictors (index) | Estimate | SE | Z-value | P-value |
|---|---|---|---|---|
| LF Gain | 3.778 | 1.649 | 2.290 | 0.022 |
| Age | -0.063 | 0.035 | -1.834 | 0.067 |
| ASPECTS/PC-ASPECTS (non-favorable) | -2.146 | 0.859 | -2.498 | 0.013 |
| Recanalization (successful) | 0.805 | 1.021 | 0.788 | 0.431 |
| Intercept: 90-day mortality | 2.831 | 2.835 | 0.999 | 0.318 |
| AIC | 61.927 | | | |
| Residual deviance | 51.927 | | | |

*AIC: Akaike information criterion.*

*Table S15. 24-hour (logistic regression) model predicting 90-day mortality.*

| Predictors (index) | Estimate | SE | Z-value | P-value |
|---|---|---|---|---|
| LF Gain | 3.106 | 1.724 | 1.802 | 0.072 |
| Age | -0.054 | 0.039 | -1.383 | 0.167 |
| 24-hour NIHSS: Moderate | -17.463 | 1854.073 | -0.009 | 0.992 |
| 24-hour NIHSS: Severe | -18.989 | 1854.073 | -0.010 | 0.992 |
| Intercept: 90-day mortality | 20.62 | 1854.075 | | |
| AIC | | | | |
| Residual deviance | | | | |

*AIC: Akaike information criterion.*
